# Treatment Outcomes and MRI Features of SUNA: A Systematic Review and Meta-Analysis

**DOI:** 10.1101/2024.08.21.24312303

**Authors:** Jay Verma, Sneha Khanduja, Mill Etienne

**Affiliations:** Maulana Azad Medical College, Delhi University; Dr. Baba Saheb Ambedkar Medical College and Hospital, Guru Gobind Singh Indraprastha University; New York Medical College

## Abstract

**Background:** Trigeminal autonomic cephalalgias (TACs), including short-lasting unilateral neuralgiform headache attacks with cranial autonomic symptoms (SUNA), are rare but debilitating headaches. The pathophysiology and optimal treatment of SUNA remain poorly defined, compounded by disparities in healthcare access.

**Objectives:** To systematically review and analyze the effectiveness of treatment options, clinical outcomes, and brain MRI findings for SUNA, and to identify gaps in the current evidence base.

**Methodology:** Following PRISMA guidelines, a systematic search was performed across multiple databases. Data from 20 studies were analyzed, focusing on treatment efficacy, patient demographics, and MRI findings. Meta-analyses were conducted on treatment effectiveness, and bias was assessed using the Newcastle-Ottawa Scale.

**Results:** Among 267 patients, the most commonly used treatments were lamotrigine (37.07%) and greater occipital nerve (GON) block (16.85%), showing effectiveness in over 50% of cases. Heterogeneity in lamotrigine effectiveness was high (Cochran’s Q = 63.10, p-value < 0.0001, α = 0.05). Lidocaine was effective for acute attacks (> 80%). Brain MRIs were mostly unremarkable, with some evidence suggesting neurovascular involvement.

**Conclusion:** Lamotrigine and GON block are effective for SUNA, though treatment responses vary widely. MRI findings often lack abnormalities, suggesting a need for further research into the pathophysiology of SUNA. Larger, high-quality studies are needed to establish standardized treatment protocols and improve patient outcomes.

## Introduction

Primary headache disorders constitute a significant global health burden, characterized by recurrent, disabling pain that can profoundly impact the quality of life of an individual.^1^ Within this heterogeneous spectrum, trigeminal autonomic cephalalgias (TACs) represent a distinct subgroup defined by the association of head pain with cranial autonomic symptoms.^2^ While TACs encompass a variety of conditions, such as cluster headaches and short-lasting unilateral neuralgiform headache attacks with cranial autonomic symptoms (SUNA), the precise pathophysiology, optimal management, and prognostic factors for these disorders remain largely elusive.^3^

SUNA, in particular, is a rare primary headache characterized by recurrent, unilateral, excruciating pain typically lasting seconds to minutes, accompanied by ipsilateral cranial autonomic features.^4^ Despite its debilitating nature, SUNA remains understudied, with a paucity of high-quality evidence to guide clinical practice. The rarity of the condition has contributed to significant knowledge gaps, hindering the development of standardized and effective treatment protocols. Consequently, patients with SUNA often experience significant delays in diagnosis and suboptimal management.^5^

Moreover, the predominance of case reports and case series in the SUNA literature underscores the limited availability of robust evidence to inform clinical decision-making. While these studies provide valuable insights into the clinical presentation and potential treatment approaches, their inherent limitations preclude definitive conclusions. A systematic review and meta-analysis of the existing literature is therefore imperative to synthesize the available evidence, identify knowledge gaps, and inform future research directions.

By comprehensively examining the treatment modalities, outcomes, and imaging features of SUNA, this study aims to contribute to the growing body of knowledge on this rare and challenging condition. Our findings could be instrumental in developing evidence-based guidelines for the diagnosis and management of SUNA, improving patient care, and reducing health disparities. Ultimately, this study seeks to enhance the understanding of SUNA and pave the way for further research to address the unmet needs of affected individuals suffering from headaches.

## Methodology

A comprehensive literature search was performed per the Preferred Reporting Items for Systematic Reviews and Meta-Analyses (PRISMA) guidelines on April 24, 2024.^6^ We used the following databases for our systematic literature review: PubMed and Google Scholar. A search strategy consisting of MeSH keywords was developed for the literature search (supplemental file S1). The survey of the databases was followed by a manual search for articles through references to the records identified from the databases. The review protocol has been a priori registered on PROSPERO (International Prospective Register of Systematic Reviews) with the protocol ID ‘CRD42024571423’, and can be accessed on the PROSPERO website.^7^

### Search Strategy and Selection Criteria

We developed a search strategy comprising MeSH keywords pertinent to our review question and the relevant patient population (supplemental file S1). Boolean operators were used during the literature search. The Rayyan software was used to manage the generated references and identify and remove duplicates. Two reviewers independently conducted the title and abstract screening, followed by the screening of full-text articles (JV and SK) (Figure 1). Any conflicts were resolved by discussion at the end of the screening process. The articles were included after screening against the following inclusion criteria: (1) included a patient population diagnosed with SUNA; (2) the diagnosis of SUNA was made according to the International Classification of Headache Disorders (ICHD-3) criteria; (3) peer-reviewed publication; (4) published in the English language; and (5) full-text was publicly accessible. Concurrently, the exclusion criteria used were: (1) conclusive diagnosis of SUNA according to the ICHD-3 criteria was not made; (2) publication not in the English language; (3) publication was not peer-reviewed; and (4) full-text was not publicly accessible. If the articles contained data from a common dataset, we included only primary patient data in our review to ensure that repetition of patient data did not occur.

**Figure 1.**
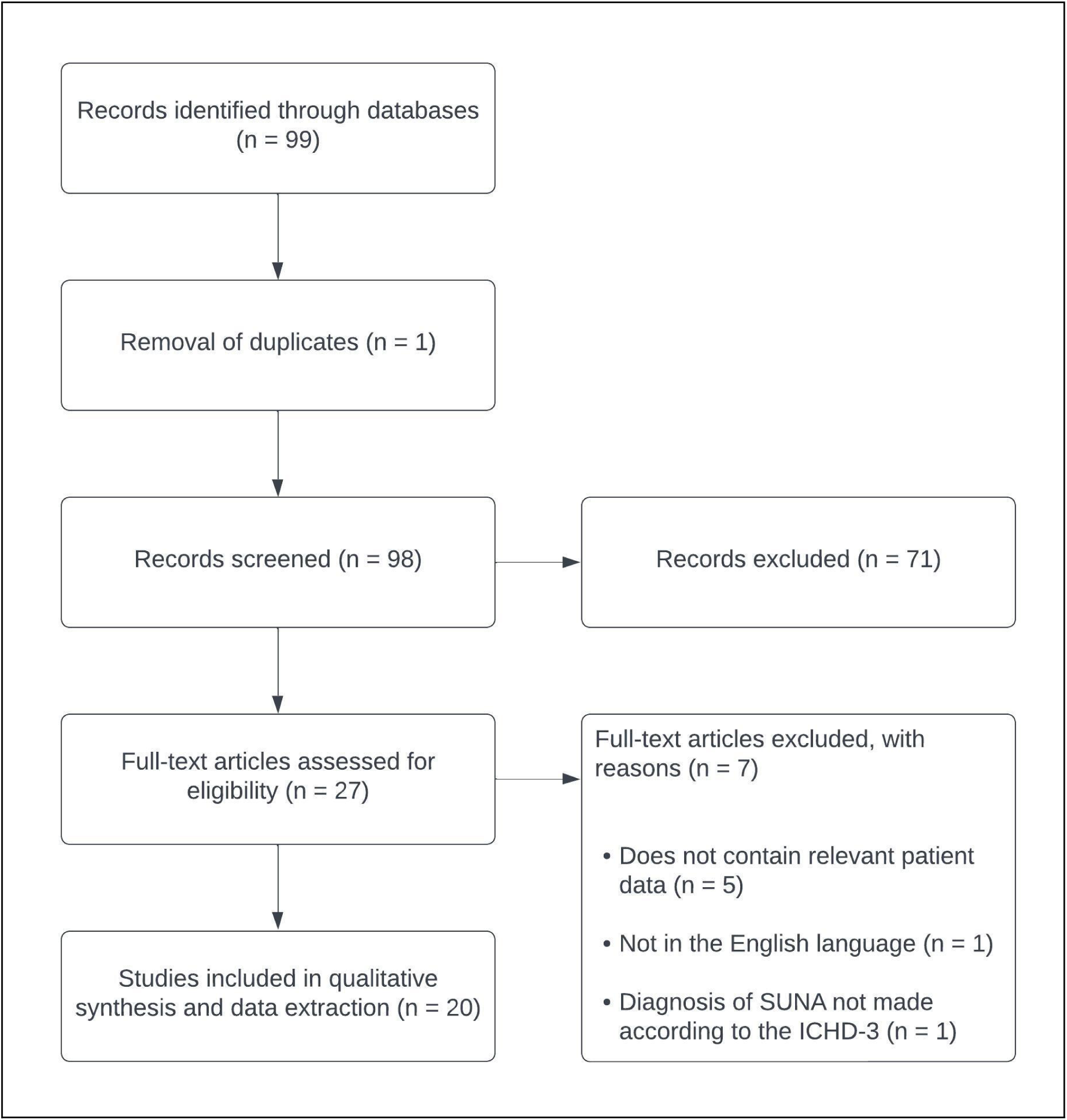
PRISMA flowchart outlining the screening and article selection process for the studies identified through PubMed and Google Scholar.

### Data Abstraction

Data abstraction was done in a tabulated form using the MS Excel software in a standard data abstraction form by two reviewers (JV and SK) (supplemental files S2 and S3). Any conflicts were resolved by an additional third reviewer (ME). We extracted data pertaining to individual patients, when available, as well as cohorts of patients. The following characteristics were considered during the data abstraction process: (1) nationality; (2) age; (3) sex; (4) brain magnetic resonance imaging (MRI) findings; (5) any diagnosed comorbidities; (6) treatment modalities with their doses; (7) duration of follow up; (8) clinical outcome; and (9) the headache attack duration and frequency before and after treatment.

### Data Synthesis and Analysis

Outcome data was generated using MS Excel after being entered in a tabulated form. We summarized variables including (1) age and sex; (2) the proportion of patients in which a certain treatment modality was used (Figure 2); (3) the proportion of patients in which a treatment modality was clinically effective, as defined by a decrease in the duration and/or frequency of the headache attacks (for the most common treatment modalities); (4) the change in headache attack frequency and duration observed with the most common treatment modalities; and (5) the correlation between a clinically effective dose of lamotrigine and the age of the patient (Figure 3). Hypothesis testing was performed using SPSS software and the chi-square test and Pearson correlation coefficient were used as applicable. The brain MRI findings extracted from our review were analyzed qualitatively and summarized by two reviewers independently to minimize bias (JV and SK). Any differences in the qualitative summaries were resolved by discussion with a third reviewer, followed by the generation of a final summary (ME).

**Figure 2.**
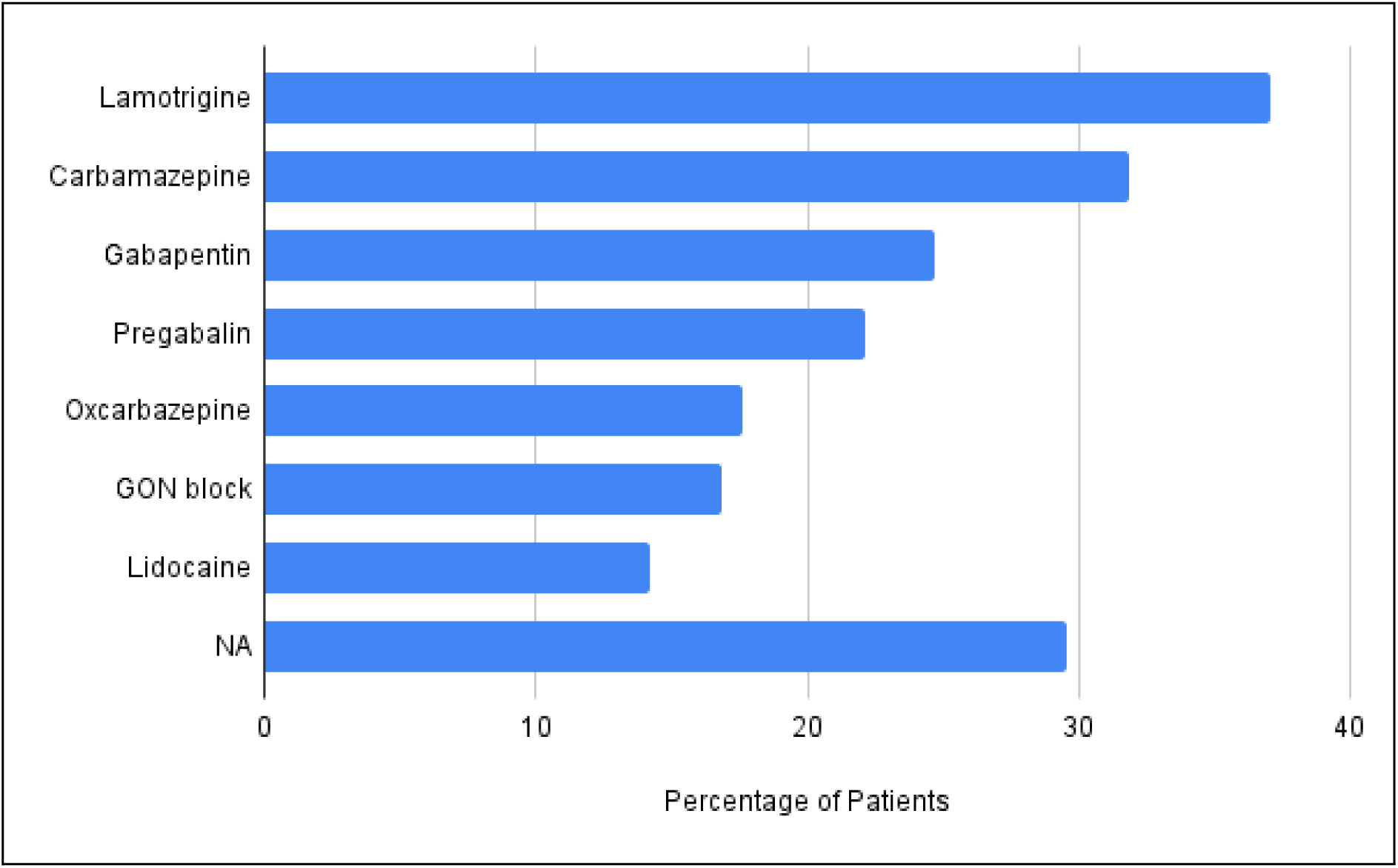
Horizontal bar graph showing the proportion of individuals in which a treatment modality was used singularly or in combination. Y-axis: seven most common treatment modalities. X-axis: percentage of patients. NA, data not available. Lamotrigine is the most frequently used preventive modality while lidocaine is the most commonly used drug for headache attack termination.

**Figure 3.**
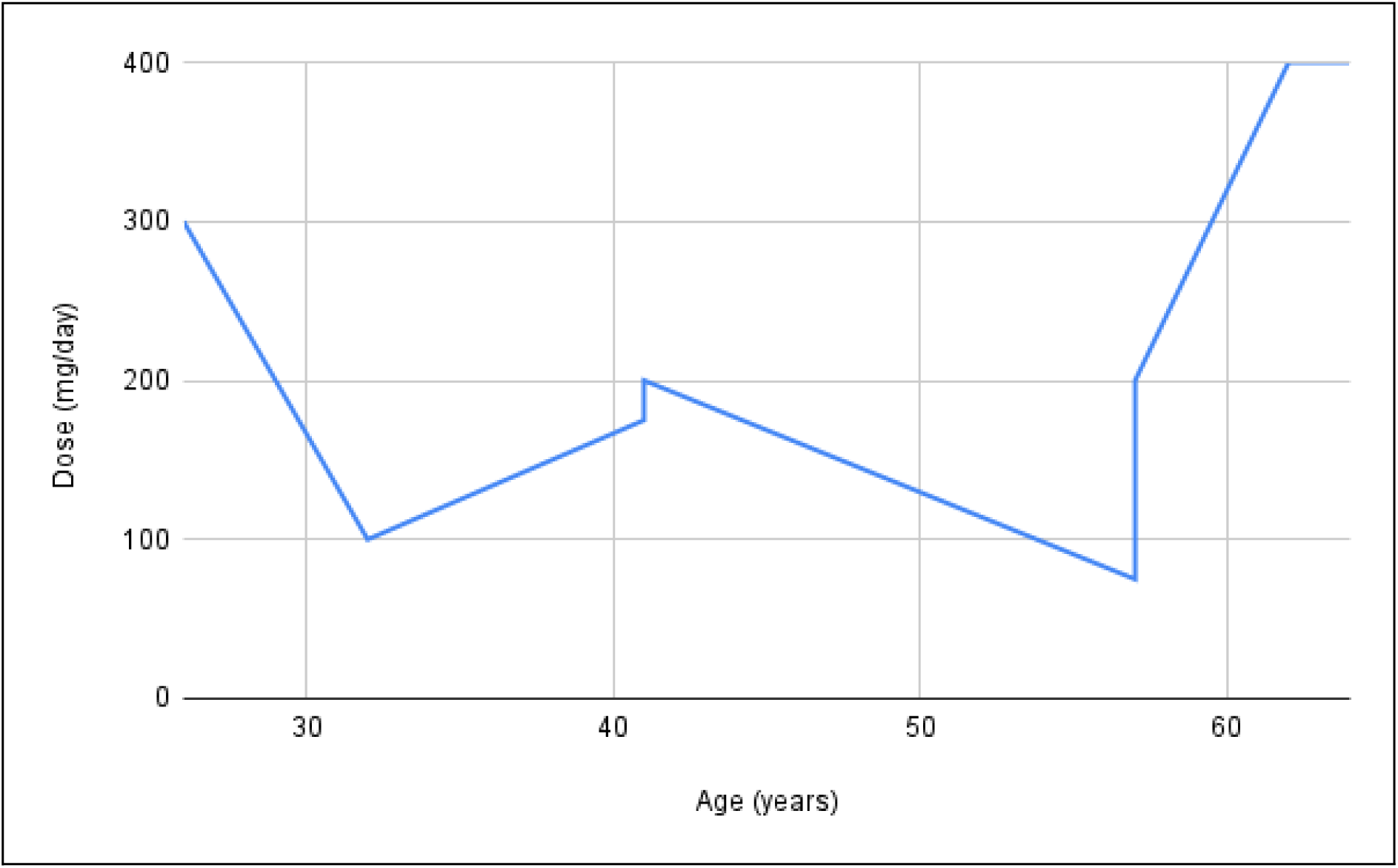
Line plot showing the correlation between a clinically effective dose of lamotrigine and age. Y-axis: clinically effective dose of lamotrigine. X-axis: age of the patient. No significant correlation exists (Pearson correlation coefficient, r = 36, r^2^ = 0.13, p-value = 0.37, α = 0.05).

We conducted a meta-analysis using a random effects model on the proportion of patients in which a treatment modality was clinically effective with a 95% confidence interval (Figures 4 to 10). Additionally, we intended to extend the meta-analysis to include the change in the frequency and duration of headache attacks as outcome measures. However, post hoc analyses revealed that these quantitative outcome measures were reported sufficiently only in case reports and case series, rendering the meta-analysis unsuitable due to none to low variation in the data (Table 2). Publication bias was assessed with a funnel plot (supplemental file S4), along with Egger’s test and Begg’s rank test. Heterogeneity was assessed using the I^2^ statistic, with I^2^ > 30%, I^2^ > 50%, and I^2^ > 75% indicating moderate, substantial, and considerable heterogeneity. Simultaneously, we also used Cochran’s Q test to evaluate heterogeneity. All analyses and figures for the meta-analysis were generated using the MedCalc software. The reporting of the meta-analysis has been done according to the Meta-analyses of Observational Studies in Epidemiology (MOOSE) checklist (supplemental file S5).^8^

**Figure 4.**
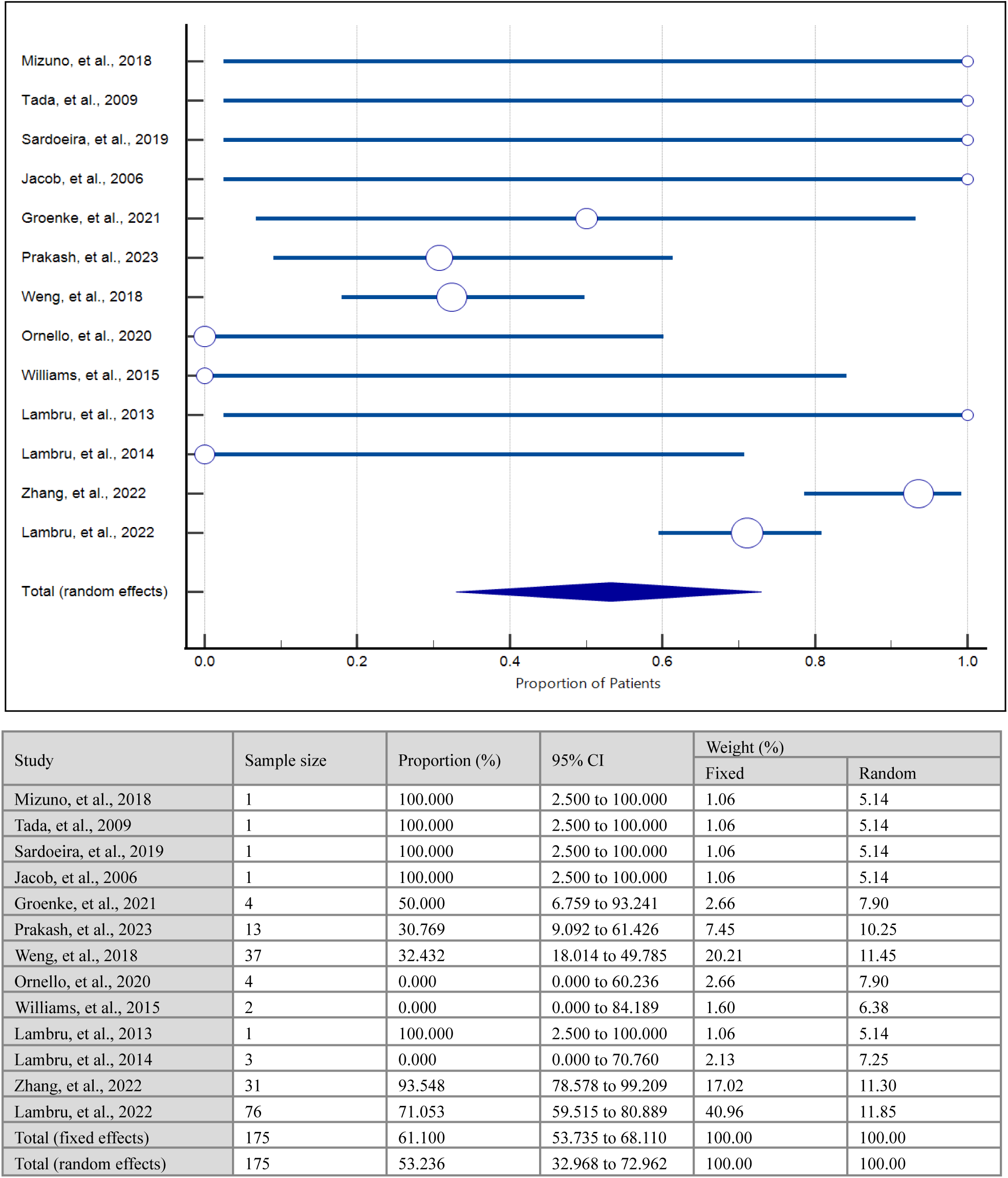
Meta-analysis: proportion of patients in whom lamotrigine was clinically effective. Cohran’s Q = 63.10, DF = 12, p-value < 0.0001, I^2^ = 80.98%, 95% CI for I^2^ = 68.48 to 88.53. Egger’s test: Intercept = −0.77, 95% CI = −3.16 to 1.61, p-value = 0.49. Begg’s test: Kendall’s tau = 0.25, p-value = 0.23.

**Figure 5.**
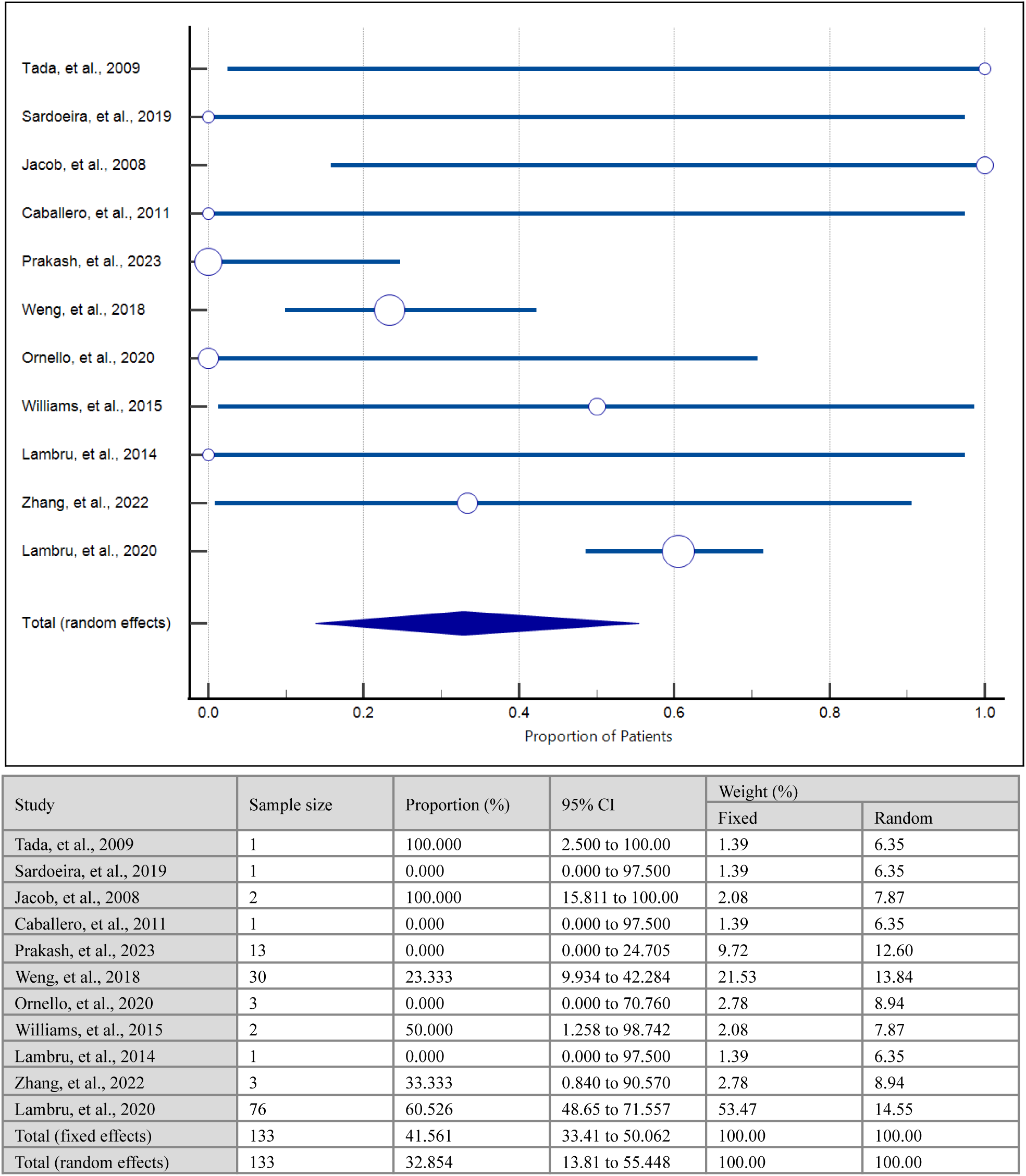
Meta-analysis: proportion of patients in whom carbamazepine was clinically effective. Cohran’s Q = 44.36, DF = 10, p-value < 0.0001, I^2^ = 77.46%, 95% CI for I^2^ = 59.86 to 87.34. Egger’s test: Intercept = −0.93, 95% CI = −3.26 to 1.40, p-value = 0.39. Begg’s test: Kendall’s tau = 0.10, p-value = 0.66.

**Figure 6.**
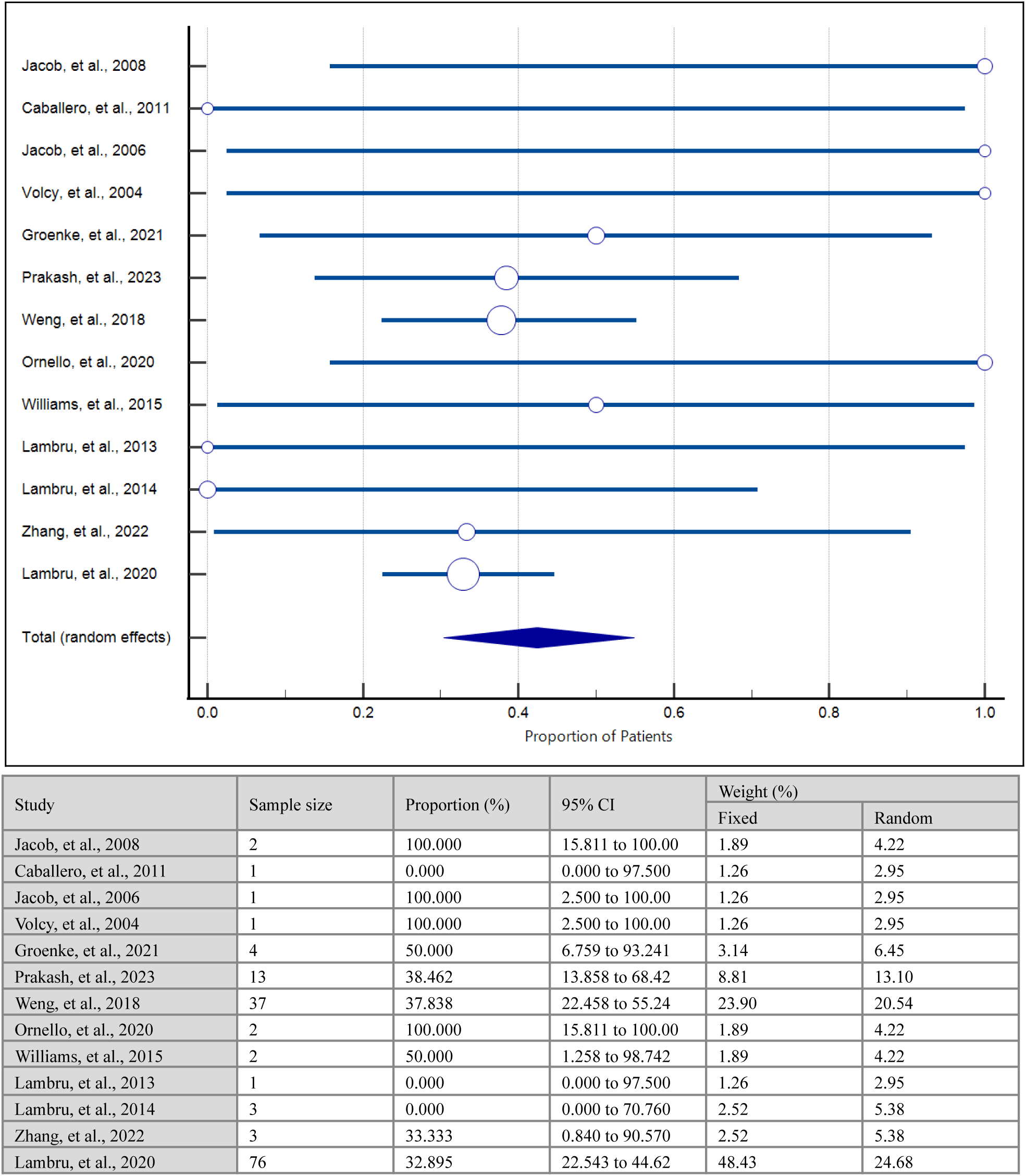
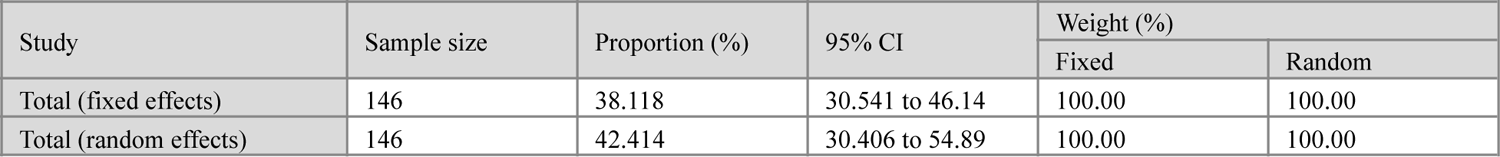
Meta-analysis: proportion of patients in whom gabapentin was clinically effective. Cohran’s Q = 17.88, DF = 12, p-value = 0.11, I^2^ = 32.91%, 95% CI for I^2^ = 0.00 to 65.32. Egger’s test: Intercept = 0.80, 95% CI = −0.34 to 1.94, p-value = 0.15. Begg’s test: Kendall’s tau = 0.22, p-value = 0.28.

**Figure 7.**
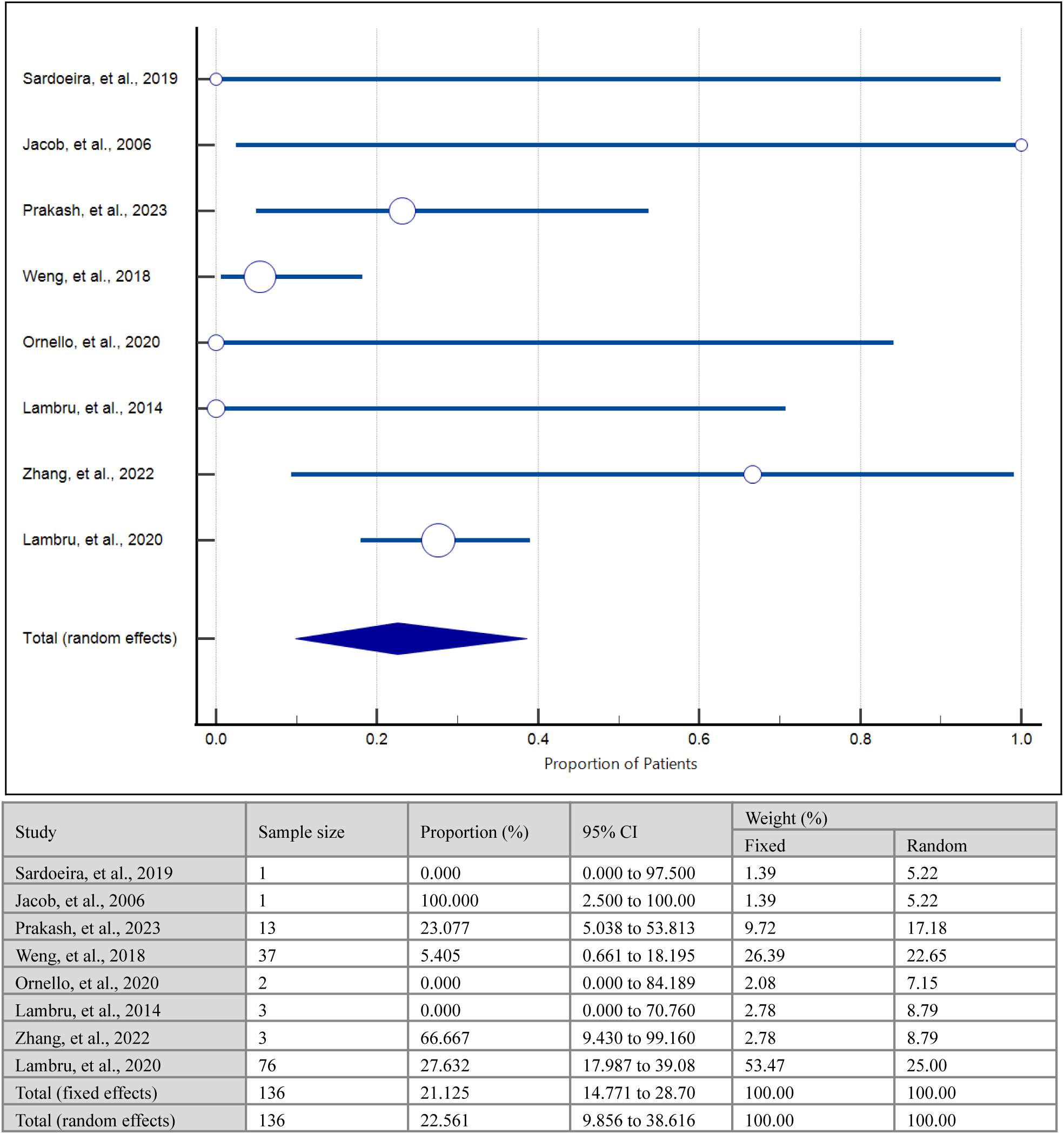
Meta-analysis: proportion of patients in whom pregabalin was clinically effective. Cohran’s Q = 17.51, DF = 7, p-value = 0.01, I^2^ = 60.03%, 95% CI for I^2^ = 13.06 to 81.63. Egger’s test: Intercept = 0.37, 95% CI = −2.07 to 2.82, p-value = 0.72. Begg’s test: Kendall’s tau = 0.07, p-value = 0.79.

**Figure 8.**
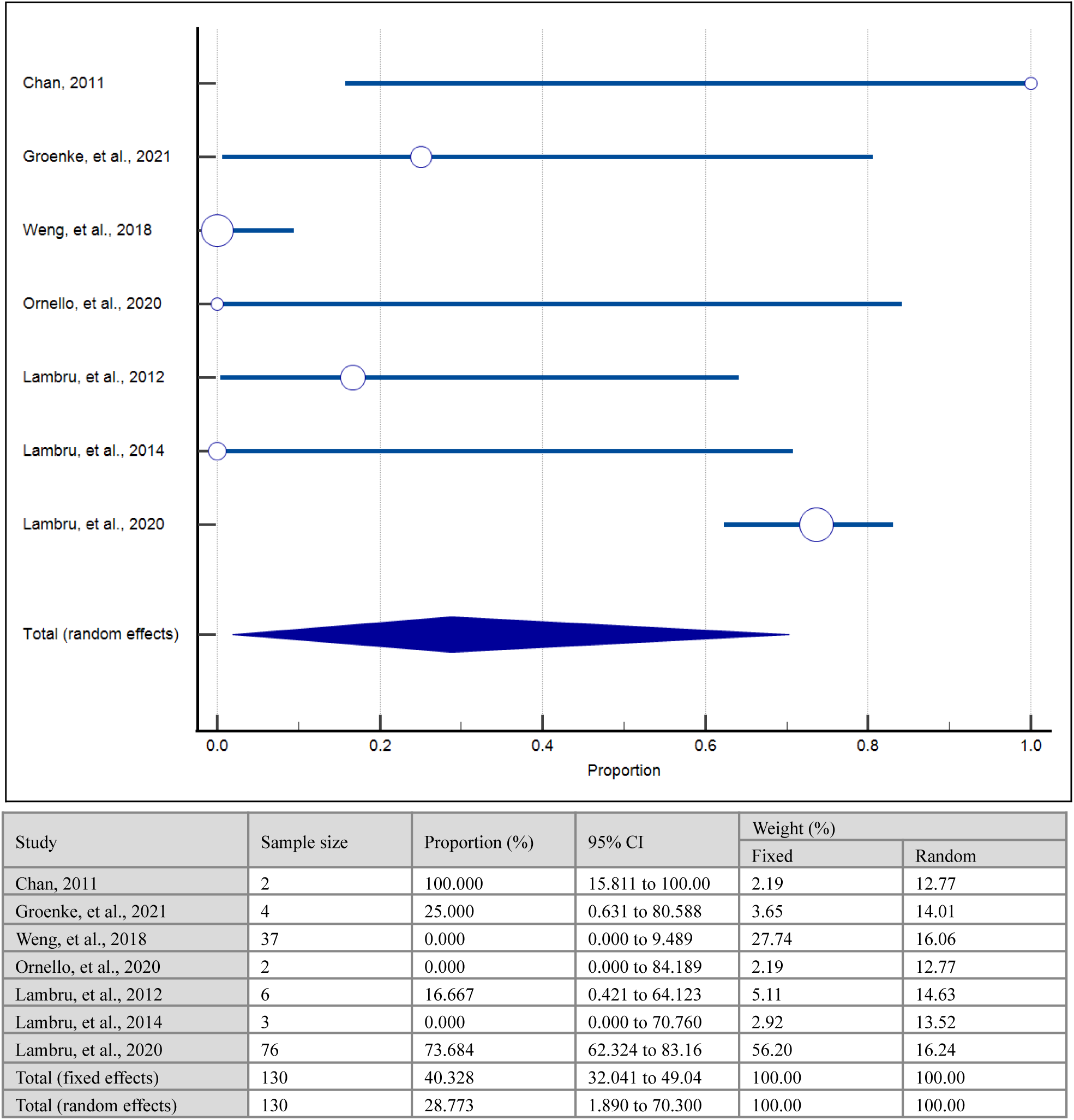
Meta-analysis: proportion of patients in whom oxcarbazepine was clinically effective. Cohran’s Q = 101.79, DF = 6, p-value < 0.0001, I^2^ = 94.11%, 95% CI for I^2^ = 90.22 to 96.45. Egger’s test: Intercept = −1.78, 95% CI = −9.09 to 5.52, p-value = 0.55. Begg’s test: Kendall’s tau = 0, p-value = 1.

**Figure 9.**
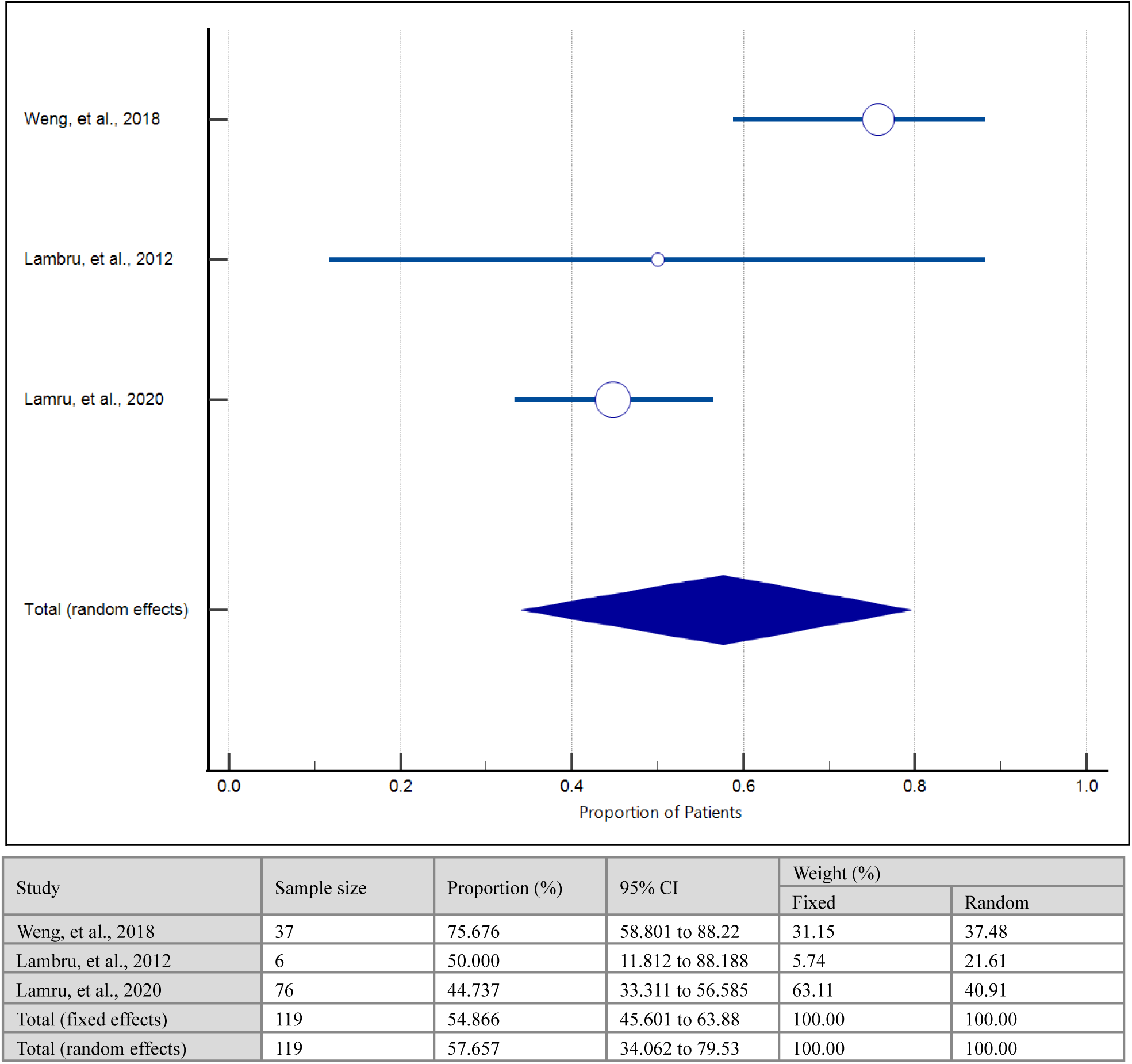
Meta-analysis: proportion of patients in whom GON block was clinically effective. Cohran’s Q = 10.10, DF = 2, p-value = 0.0064, I^2^ = 80.21%, 95% CI for I^2^ = 37.55 to 93.73. Egger’s test: Intercept = 1.29, 95% CI = −55.59 to 58.17, p-value = 0.82. Begg’s test: Kendall’s tau = 0.33, p-value = 0.60.

**Figure 10.**
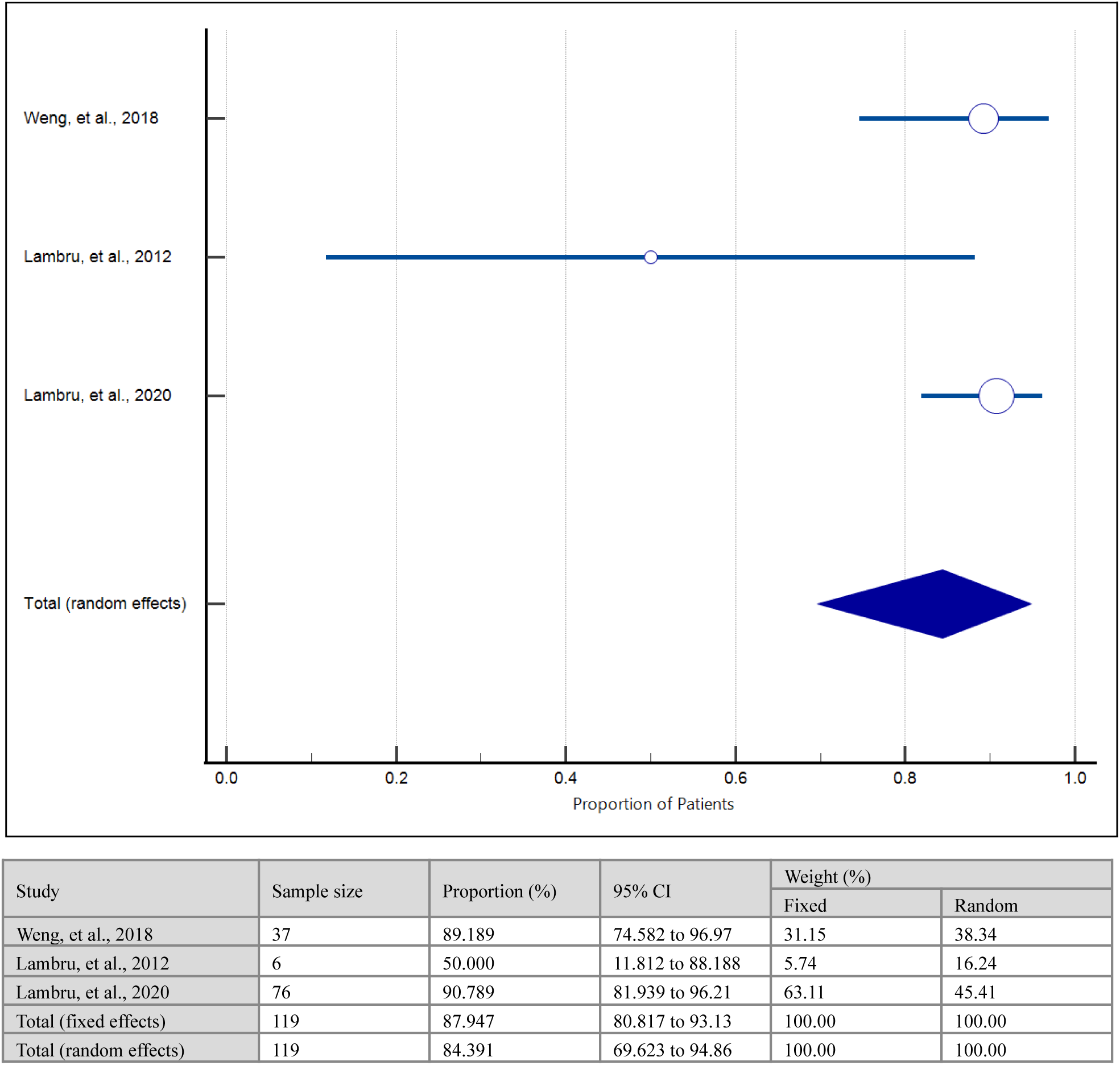
Meta-analysis: proportion of patients in whom lidocaine was clinically effective. Cohran’s Q = 5.63, DF = 2, p-value = 0.05, I^2^ = 64.47%, 95% CI for I^2^ = 0.00 to 89.81. Egger’s test: Intercept = −3.39, 95% CI = −13.10 to 6.32, p-value = 0.14. Begg’s test: Kendall’s tau = −1.00, p-value = 0.11.

### Risk of Bias Analysis

Since the majority of studies included in the review were cohort studies and case reports or case series, the Newcastle-Ottawa scale (NOS) for cohort studies and case reports was developed and used for quality appraisal. Two reviewers (JV and SK) independently assessed the risk of bias. Any conflicts in the scoring were resolved by discussion with a third reviewer (ME). The NOS was constructed to evaluate the quality of studies according to the following domains: (1) study design; (2) patient population; (3) content of the study; and (4) reporting of outcomes such that the data could be incorporated into our review. After the NOS was developed, the studies were scored against three categories: (1) selection (four subcategories, four stars maximum); (2) comparability (one subcategory, two stars maximum); and (3) outcome (two subcategories, three stars maximum). The full list of adapted categories and subcategories from the NOS is contained in supplemental file S6 along with the results of the risk of bias analysis.

### Patient Involvement and Ethical Considerations

No patients were involved in this study and no retrospective patient data was collected from the institutes to which the authors are affiliated.

## Results

### Literature Search and Screening Results

The systematic literature review yielded a total of n = 99 studies from the following databases: PubMed and Google Scholar. After the removal of one duplicate, n = 98 articles were included in the title and abstract screening to determine the relevance to our review question. 71 articles were, thus, excluded at this stage because those either did not report treatment outcomes and/or brain MRI findings or did not include patients with an ICHD-3 diagnosis of SUNA. After this step, full-text screening was performed on n = 27 articles, of which seven were excluded, with their reasons summarized in Figure 1. Finally, we included n = 20 articles in our review.

### Study Characteristics

The majority of studies included in our review were case reports and case series with two cohort studies, one cross-sectional study, and one randomized control trial. The sample size ranged from one to 79 participants. Table 1 contains a summary of all of the 20 articles. Furthermore, nine studies were conducted in the United Kingdom (UK), three in the United States of America (USA), and the rest across Asian and European countries such as India (n = 1), China (n = 1), Denmark (n = 1), Portugal (n = 1), Spain (n = 1), and Japan (n = 2) with one article from Australia.

**Table 1.**
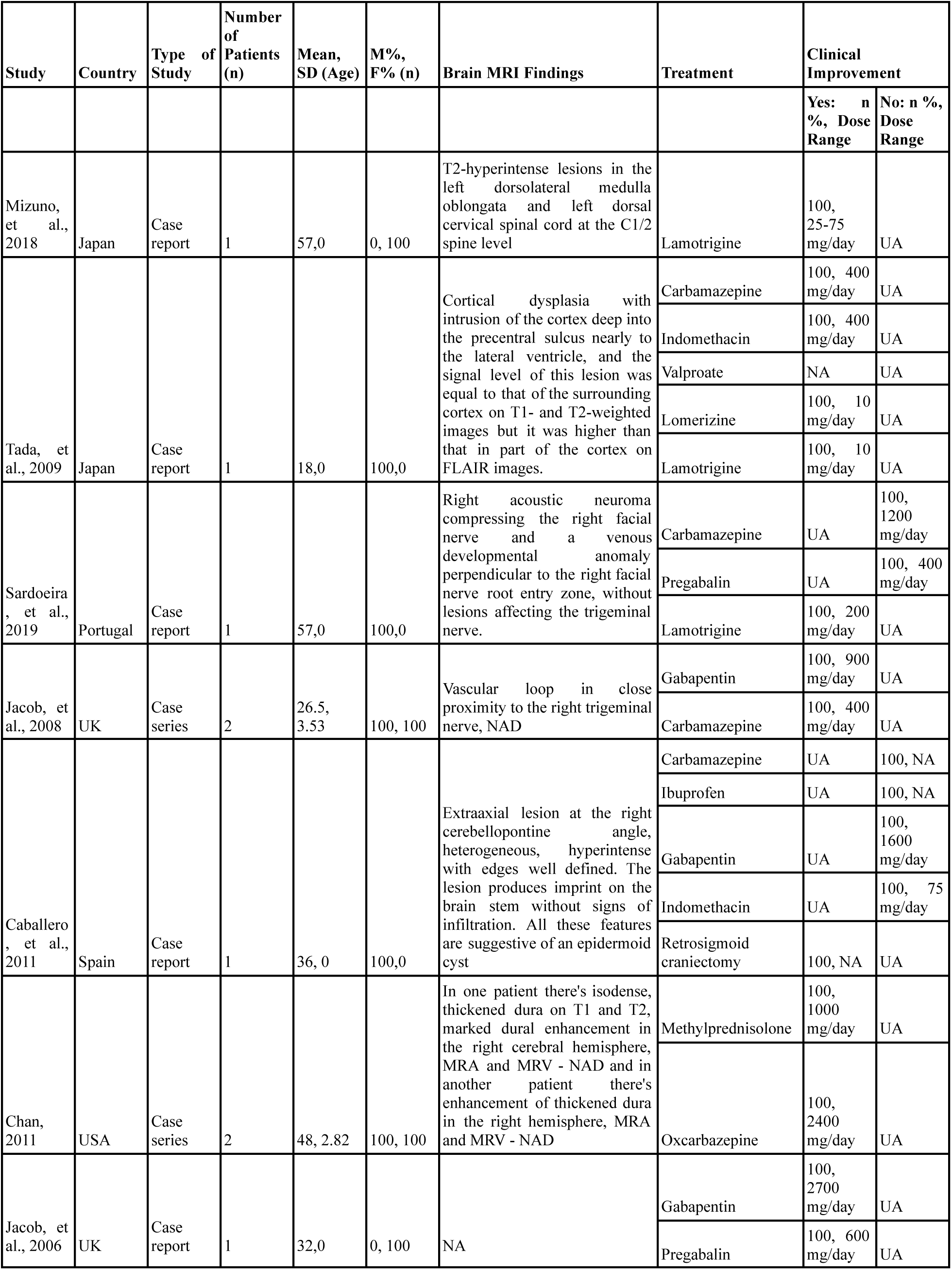
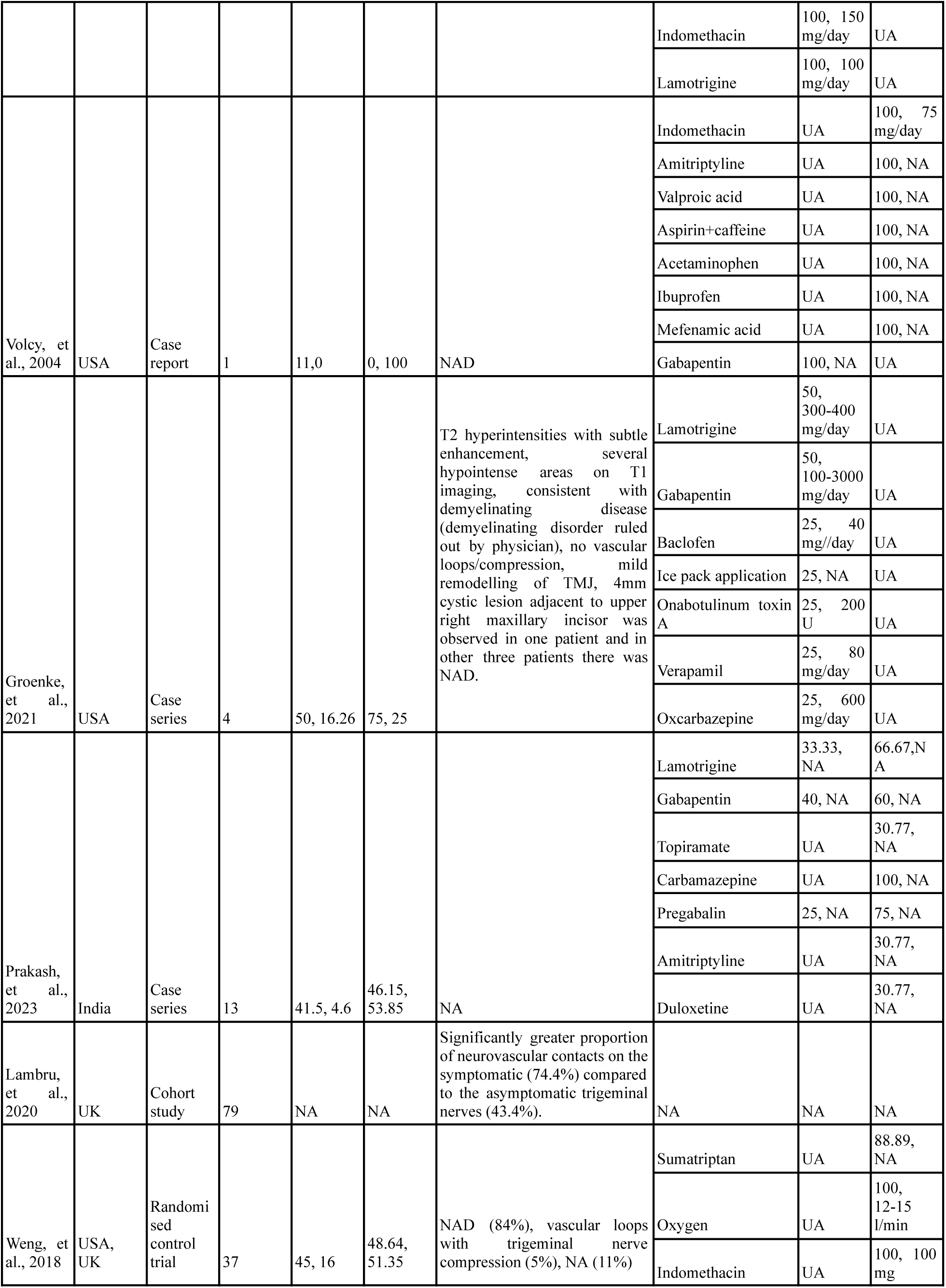
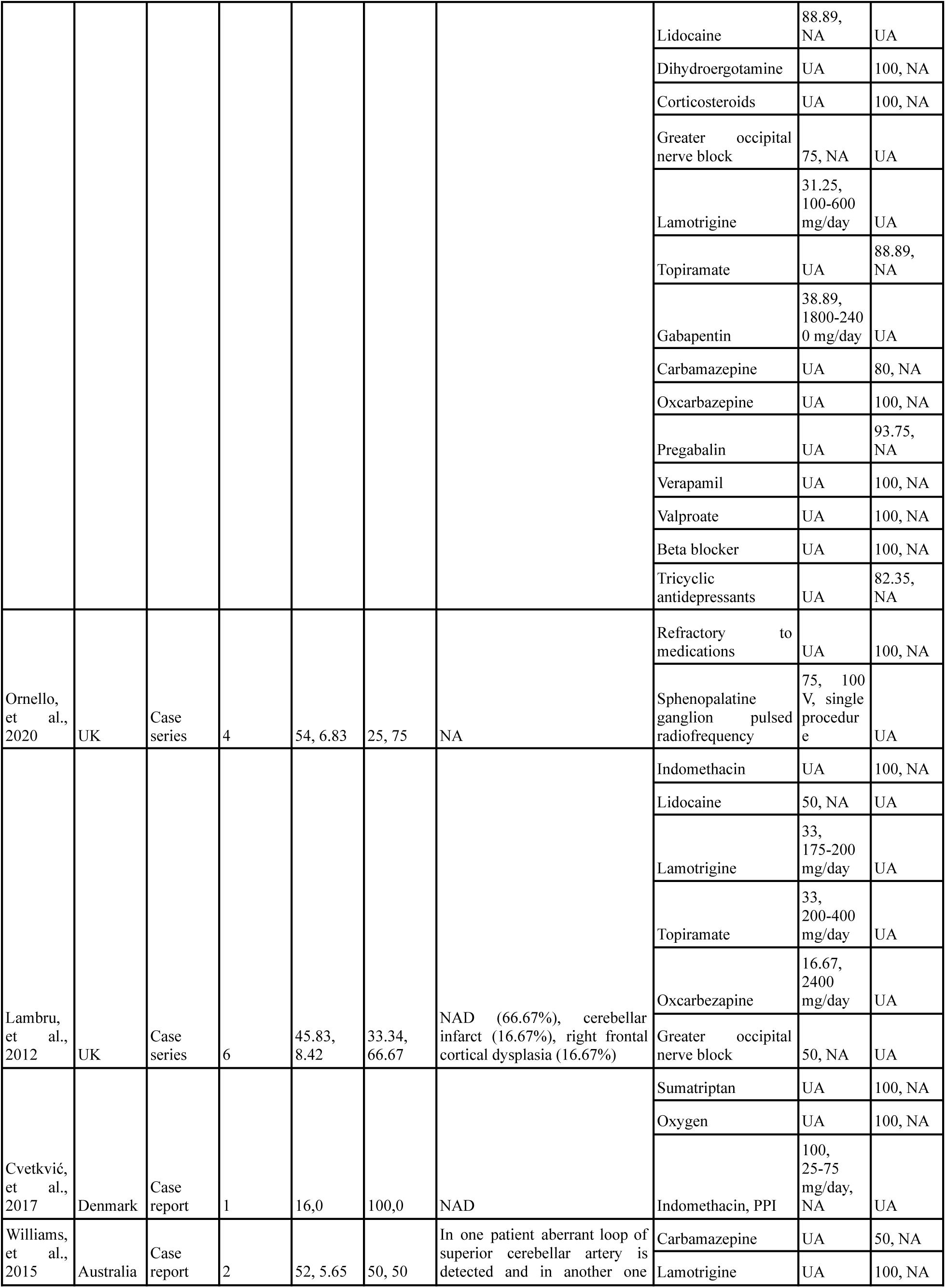
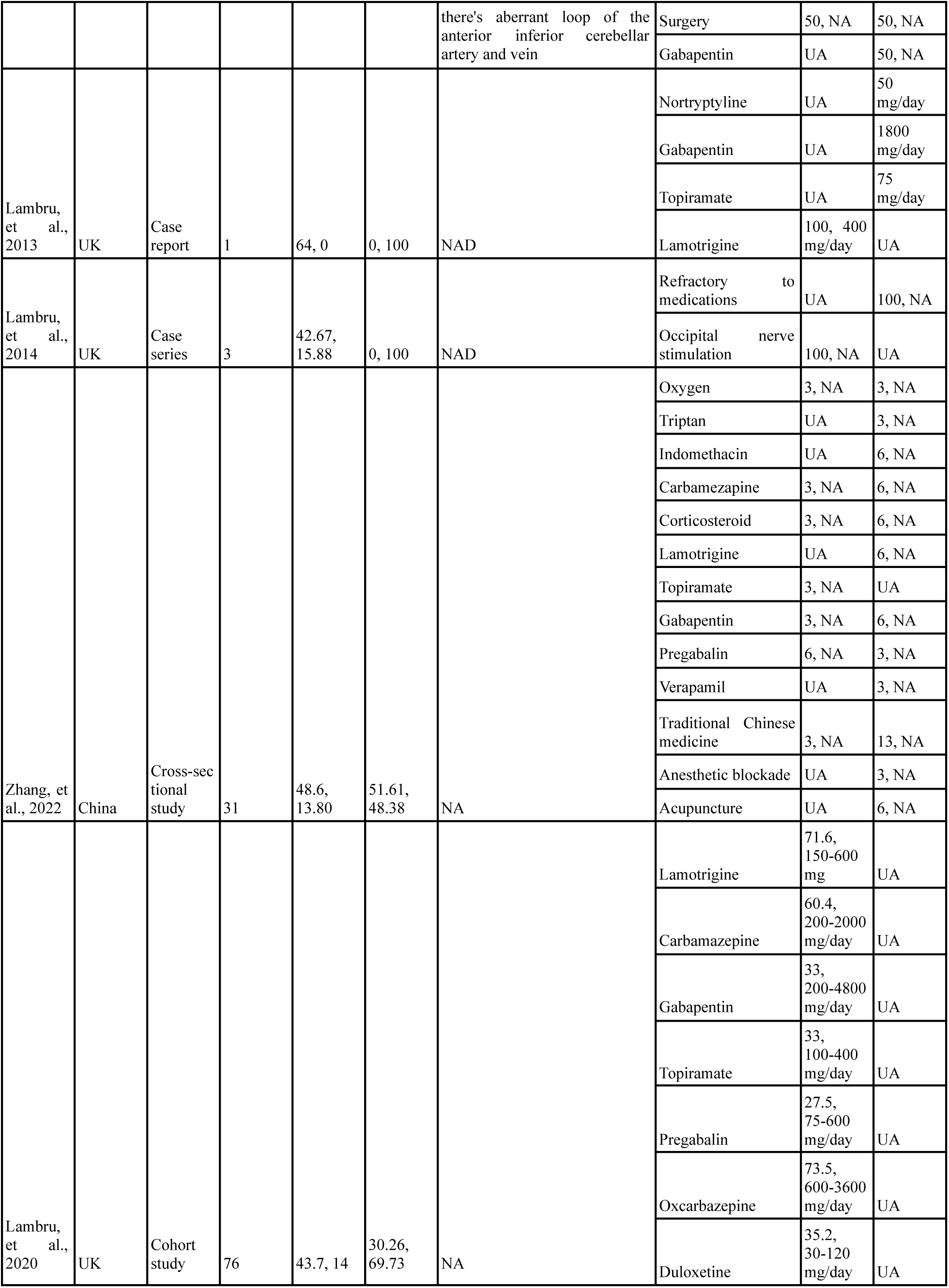
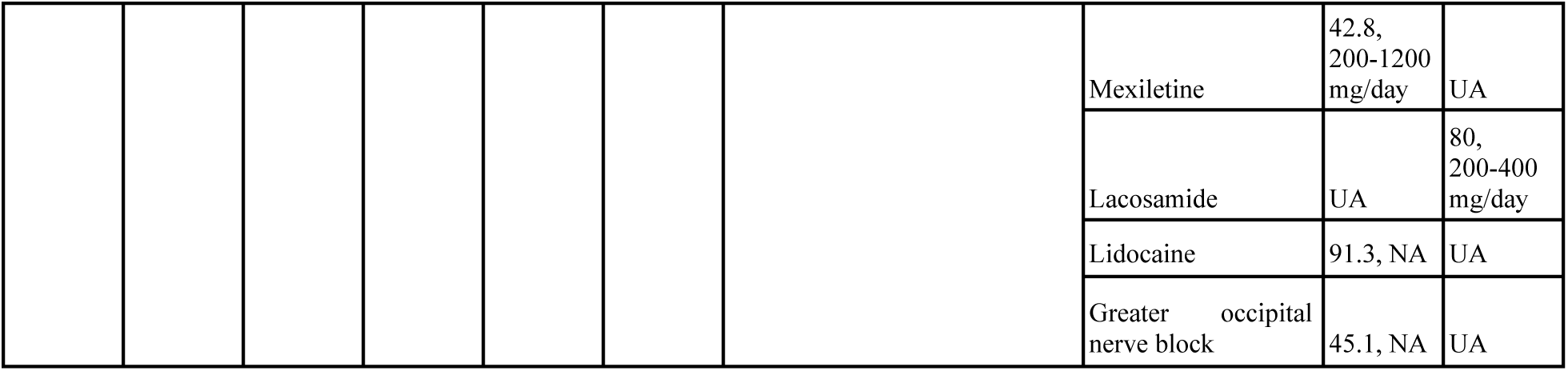
Study characteristics. NA, data not available. UA, not applicable. NAD, no abnormality detected. SD, standard deviation. M%, percentage of male patients. F%, percentage of female patients.

**Table 2.**
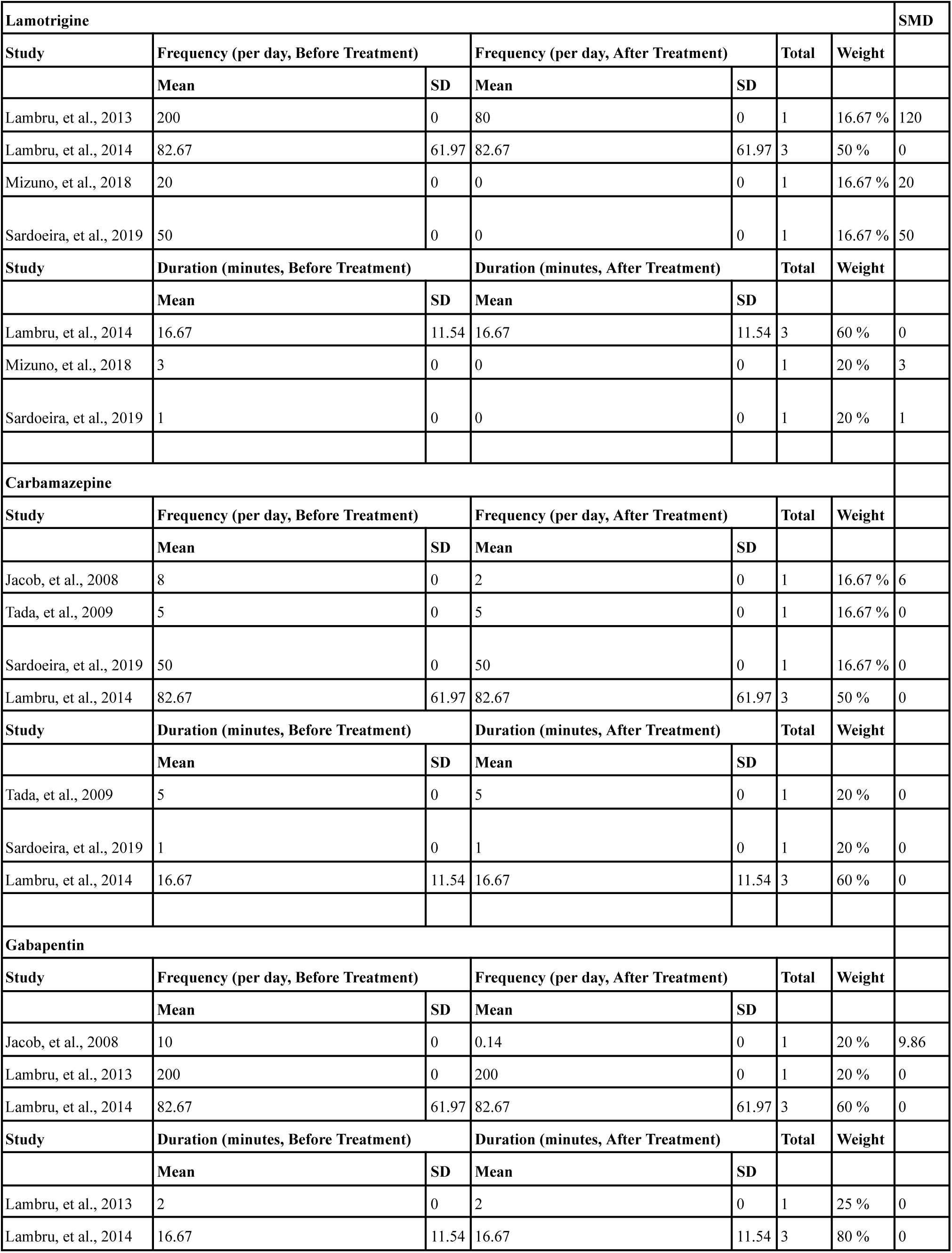
Headache attack duration and frequency data, before and after treatment, for selected treatment modalities.

### Risk of Bias Analysis

Most studies included in this review were case reports and case series (n = 16). Supplemental file S6 contains the complete NOS results for all studies assessed for bias, determined using the NOS protocol detailed in the Methodology. Most studies were moderate in quality.

### Demographic Characteristics

We obtained individual participant data from a total of 267 patients, from which subgroups were chosen for various analyses depending on the availability of the data. The overall study population had a mean age of 31.46 years with a standard deviation of 13.92 years. In terms of gender distribution, 26.96% of the patients were male and 43.46% were female, demonstrating a strong female predominance (*χ*^2^ = 5.62, p-value = 0.01, α = 0.05), which is in line with previous literature.^9^

### Treatment Modalities and Outcomes

A wide range of treatment options were used across various studies included in our review, ranging from non-steroidal anti-inflammatory drugs (NSAIDs) and steroids, to anti-epileptic medications like valproate and medications for neuropathic pain such as carbamazepine, gabapentin, and pregabalin. The most common treatment modality, however, is lamotrigine (37.07%). Surgical options included greater occipital nerve (GON) block (16.85%) and sphenopalatine ganglion pulsed radiofrequency. The usage frequency of the most common treatment options is shown in Figure 2, while a comprehensive list of all the treatment methods used in all patients is contained in Table 1.

Previous literature has shown that an effective dose of lamotrigine may vary with age due to pharmacokinetic changes associated with aging. To test this theory on clinical data, we determined the correlation between the age of the patients and a clinically effective dose of lamotrigine (Figure 3).^10^ Our analysis demonstrated that no such significant correlation existed (r = 36, r^2^ = 0.13, p-value = 0.37, α = 0.05), although the number of participants included in this analysis was small (n = 8). For comprehensiveness, we attempted to investigate the correlation between age and a clinically ineffective dose of lamotrigine. However, data on dosage in patients where lamotrigine was not clinically effective was not reported in quantities sufficient to calculate a Pearson correlation coefficient.

Finally, we proceeded to calculate the proportion of patients in which a treatment modality was clinically effective, as described in the Methodology. The results of the meta-analyses are shown in Figures 4 to 10. The most effective preventive treatment options included lamotrigine and GON block with effectiveness slightly higher than 50%, whereas the effectiveness of the rest of the preventive methods was restricted to less than 50%. Of note, in the studies sampled for lamotrigine effectiveness significant heterogeneity existed (Cochran’s Q = 63.10, DF = 12, p-value < 0.0001, α = 0.05, I^2^ = 80.98%, 95% CI = 68.48 to 88.53). Moreover, the number of studies in which the GON block was used was quite less (n = 3), resulting in a relatively small sample size in terms of individual participants (n = 45). Among therapeutic treatment modalities, lidocaine was both the most commonly used (14.23%) and the most effective (> 80%) modality. Although the number of studies sampled for lidocaine effectiveness was small (n = 3), they demonstrated statistically insignificant publication bias and heterogeneity. To investigate patient outcomes more quantitatively, we abstracted data pertaining to the duration and frequency of headache attacks before and after treatment, when available (supplemental file S3). Since most of the studies that contained these data were case reports or case series, no to low standard deviation was observed, rendering meta-analyses unsuitable. A summary of the headache attack duration and frequency data along with standardized mean differences is presented in Table 2.

### Brain MRI Findings

For the majority of patients, brain MRI findings were not reported (n = 125, 46.81%), and for a significant proportion of patients (n = 45, 16.85%), brain MRI did not capture any abnormality associated with SUNA. Two studies specifically assessed the correlation between SUNA and trigeminal nerve compression, as determined by the presence of neurovascular contacts (Weng, et al., 2018 and Lambru, et al., 2020).^11,12^ Lambru, et al., 2018 reported a significantly greater proportion of neurovascular contacts on symptomatic trigeminal nerves (74.4%) compared to asymptomatic trigeminal nerves (43.4%), whereas Weng, et al., 2018 reported trigeminal nerve compression in only two out of 37 patients, with no abnormality being detected in 31 patients. Another case series (Jacob, et al., 2008) reported close proximity between a vascular loop and the trigeminal nerve in one out of two patients, further suggesting that the pathophysiology of SUNA might be linked to trigeminal nerve compression by vascular structures.^13^ A full list of all of the brain MRI findings from the studies included in our review is contained in Table 1.

## Discussion

This systematic review and meta-analysis provide a comprehensive evaluation of the treatment modalities, outcomes, and brain MRI findings associated with SUNA. Our findings contribute valuable insights into this rare and challenging condition, highlighting several key areas of interest and identifying both the strengths and limitations of the current literature. A proposed management guideline for SUNA, based on our findings, is presented in Figure 11.

**Figure 11.**
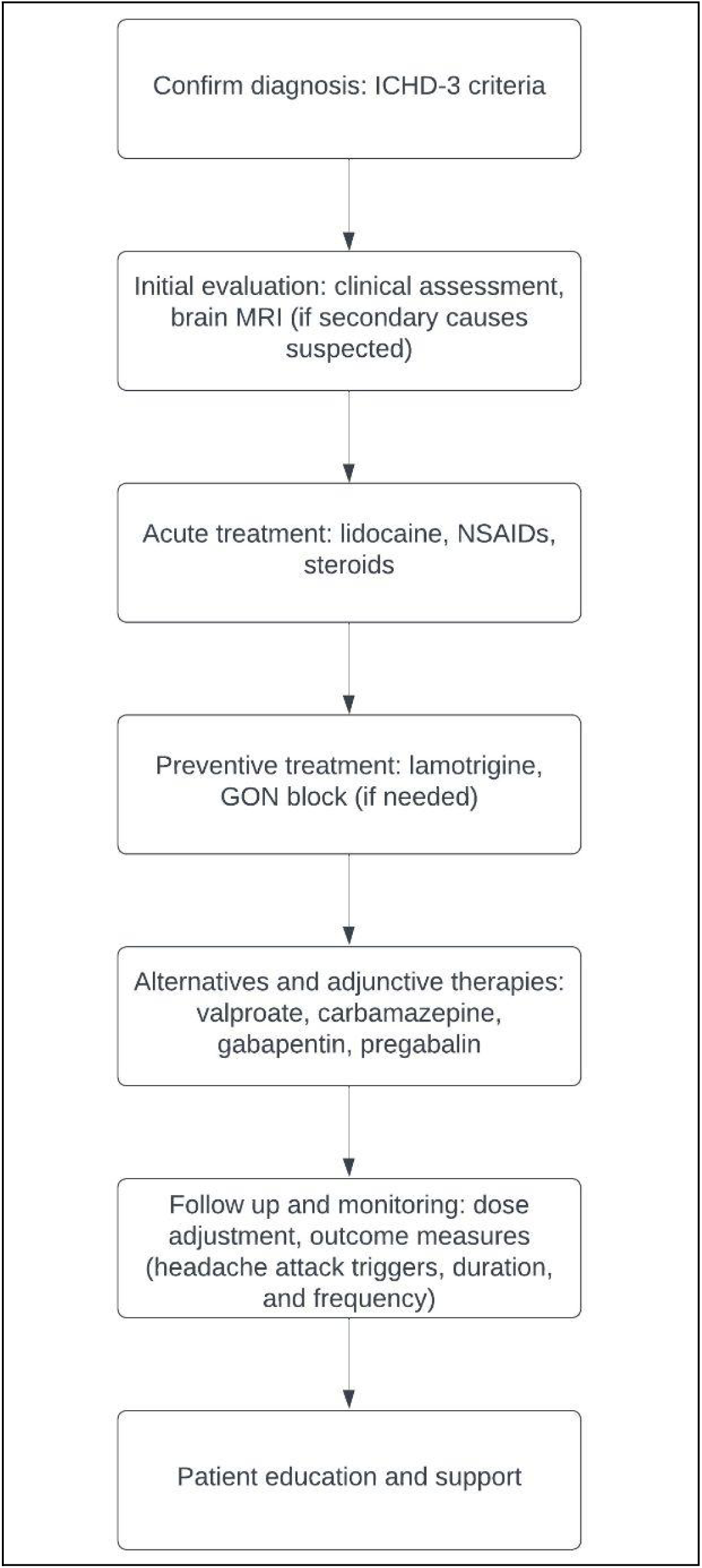
Proposed management guideline for SUNA, based on our systematic review and meta-analysis.

### Treatment Modalities and Their Effectiveness

Our analysis reveals a broad range of treatment options utilized for SUNA, including both pharmacological and procedural interventions. Lamotrigine and greater occipital nerve (GON) block emerged as the most commonly used and potentially effective treatments, with a clinical effectiveness rate slightly above 50%. This aligns with the previous evidence suggesting that lamotrigine may offer relief for some patients, though the variability in effectiveness underscores the need for more targeted research.^14^ The high heterogeneity observed in the effectiveness of lamotrigine (I² = 80.98%) suggests substantial variation in treatment responses across studies. This could be attributed to differences in dosing regimens, patient demographics, and methodological variations among studies.^15^

Lidocaine was found to be a highly effective therapeutic option for acute attacks, aligning with its established role in managing other trigeminal neuralgias.^16^ The low standard deviation in headache frequency and duration data suggests that, while lidocaine may be effective, further research with larger sample sizes is needed to confirm these results and assess its long-term impact.

The lack of a significant correlation between age and the effective dose of lamotrigine indicates that age may not be a critical factor in determining dosage efficacy. However, the small sample size for this analysis limits the robustness of this conclusion. This highlights the need for larger, well-designed studies to better understand the pharmacokinetics and dosing strategies for lamotrigine in SUNA patients.^17–20^

### Brain MRI Findings

Our review found that brain MRI results were often unremarkable, with no abnormalities detected in a significant proportion of patients. This aligns with the notion that SUNA may not be associated with visible structural abnormalities in many cases.^21^ However, the presence of neurovascular contacts or trigeminal nerve compression in some studies suggests a potential pathophysiological link between vascular structures and trigeminal nerve irritation.^22^ The conflicting findings from studies on trigeminal nerve compression further emphasize the need for more focused research in this area. Understanding the underlying neurovascular relationships could provide critical insights into the pathogenesis of SUNA and guide the development of more effective treatments.^23^

### Study Limitations and Implications

A major limitation of this review is the predominance of case reports and case series among the included studies, which inherently limits the generalizability and reliability of the findings. The moderate quality of the included studies, as indicated by the risk of bias analysis, further suggests that the evidence base for SUNA remains limited and fragmented. The small sample sizes and variability in study methodologies contribute to the challenges of drawing definitive conclusions.

Despite these limitations, this review underscores the importance of developing standardized treatment protocols for SUNA and encourages further research into both pharmacological and procedural interventions.^24^ Future studies should aim to include larger, multicenter cohorts and employ rigorous methodologies to enhance the quality of evidence. Additionally, exploring patient-reported outcomes and quality-of-life measures could provide a more comprehensive understanding of the impact of SUNA and its treatments.^11^

## Conclusions

In summary, this systematic review and meta-analysis reveal that lamotrigine and GON block are among the most commonly used and effective treatments for SUNA, though significant variability in treatment response and study methodologies underscores the need for more personalized and rigorous research. Brain MRI findings often show no abnormalities, suggesting that SUNA may not be linked to visible structural changes, yet some evidence of neurovascular interactions warrants further investigation into its pathophysiology. The predominance of case reports and moderate-quality studies highlights the need for larger, well-designed trials to establish standardized treatment protocols and enhance our understanding of SUNA.

## Data Availability

All data produced in the present study are available upon reasonable request to the authors.

## Acknowledgments

The authors of this study duly acknowledge the contributions of Pulkit Jain, MBBS (Maulana Azad Medical College, University of Delhi) and Pratischtha Kain, MBBS (Maulana Azad Medical College, University of Delhi) in the full-text screening of articles and abstraction of data from the selected articles.

## References

1. Kopel, D., & Gottschalk, C. (2022). The Epidemiology of Primary Headache Disorders. Seminars in neurology, 42(4), 449–458. 10.1055/a-1942-6823

2. Benoliel R. (2012). Trigeminal autonomic cephalgias. British journal of pain, 6(3), 106–123. 10.1177/2049463712456355

3. Lambru, G., & Matharu, M. S. (2012). Trigeminal autonomic cephalalgias: A review of recent diagnostic, therapeutic and pathophysiological developments. Annals of Indian Academy of Neurology, 15(Suppl 1), S51–S61. 10.4103/0972-2327.100007

4. Pomeroy, J. L., & Nahas, S. J. (2015). SUNCT/SUNA: A Review. Current pain and headache reports, 19(8), 38. 10.1007/s11916-015-0511-2

5. Torres-Romero, C. M., Romaña-Espiritu, P., Macías-de la Cruz, J. H., & Sauri-Suárez, S. (2023). SUNCT/SUNA: ¿frecuentemente mal diagnosticada como neuralgia del trigémino? [SUNCT/SUNA: frequently misdiagnosed as trigeminal neuralgia?]. Revista de neurologia, 77(2), 41–46. 10.33588/rn.7702.2023166

6. Page, M. J., McKenzie, J. E., Bossuyt, P. M., Boutron, I., Hoffmann, T. C., Mulrow, C. D., Shamseer, L., Tetzlaff, J. M., Akl, E. A., Brennan, S. E., Chou, R., Glanville, J., Grimshaw, J. M., Hróbjartsson, A., Lalu, M. M., Li, T., Loder, E. W., Mayo-Wilson, E., McDonald, S., McGuinness, L. A., … Moher, D. (2021). The PRISMA 2020 statement: an updated guideline for reporting systematic reviews. Systematic reviews, 10(1), 89. 10.1186/s13643-021-01626-4

7. Schiavo J. H. (2019). PROSPERO: An International Register of Systematic Review Protocols. Medical reference services quarterly, 38(2), 171–180. 10.1080/02763869.2019.1588072

8. Stroup, D. F., Berlin, J. A., Morton, S. C., Olkin, I., Williamson, G. D., Rennie, D., Moher, D., Becker, B. J., Sipe, T. A., & Thacker, S. B. (2000). Meta-analysis of observational studies in epidemiology: a proposal for reporting. Meta-analysis Of Observational Studies in Epidemiology (MOOSE) group. JAMA, 283(15), 2008–2012. 10.1001/jama.283.15.2008

9. Favoni, V., Grimaldi, D., Pierangeli, G., Cortelli, P., & Cevoli, S. (2013). SUNCT/SUNA and neurovascular compression: new cases and critical literature review. Cephalalgia: an international journal of headache, 33(16), 1337–1348. 10.1177/0333102413494273

10. Reimers, A., Skogvoll, E., Sund, J. K., & Spigset, O. (2007). Lamotrigine in children and adolescents: the impact of age on its serum concentrations and on the extent of drug interactions. European journal of clinical pharmacology, 63(7), 687–692. 10.1007/s00228-007-0308-2

11. Weng, H. Y., Cohen, A. S., Schankin, C., & Goadsby, P. J. (2018). Phenotypic and treatment outcome data on SUNCT and SUNA, including a randomised placebo-controlled trial. Cephalalgia: an international journal of headache, 38(9), 1554–1563. 10.1177/0333102417739304

12. Lambru, G., Stubberud, A., Rantell, K., Lagrata, S., Tronvik, E., & Matharu, M. S. (2021). Medical treatment of SUNCT and SUNA: a prospective open-label study including single-arm meta-analysis. Journal of neurology, neurosurgery, and psychiatry, 92(3), 233–241. 10.1136/jnnp-2020-323999

13. Jacob, S., Saha, A., & Rajabally, Y. (2008). Post-traumatic short-lasting unilateral headache with cranial autonomic symptoms (SUNA). Cephalalgia: an international journal of headache, 28(9), 991–993. 10.1111/j.1468-2982.2008.01622.x

14. Lambru, G., Stubberud, A., Rantell, K., Lagrata, S., Tronvik, E., & Matharu, M. S. (2021). Medical treatment of SUNCT and SUNA: a prospective open-label study including single-arm meta-analysis. Journal of neurology, neurosurgery, and psychiatry, 92(3), 233–241. 10.1136/jnnp-2020-323999

15. Pareja, J. A., Alvarez, M., & Montojo, T. (2013). SUNCT and SUNA: Recognition and Treatment. Current treatment options in neurology, 15(1), 28–39. 10.1007/s11940-012-0211-8

16. Han, K. R., Kim, C., Chae, Y. J., & Kim, D. W. (2008). Efficacy and safety of high concentration lidocaine for trigeminal nerve block in patients with trigeminal neuralgia. International journal of clinical practice, 62(2), 248–254. 10.1111/j.1742-1241.2007.01568.x

17. Shawahna, R., Saba’aneh, H., Daraghmeh, A., Qassarwi, Y., Franco, V., & Declèves, X. (2023). Solubility of lamotrigine in age-specific biorelevant media that simulated the fasted- and fed-conditions of the gastric and intestinal environments in pediatrics and adults: implications for traditional, re-formulated, modified, and new oral formulations. BMC biotechnology, 23(1), 36. 10.1186/s12896-023-00809-2

18. Wang, M. L., Wang, H. X., Zhao, M. M., Ma, Y. Y., & Zhao, L. M. (2021). Redefining the age-specific therapeutic ranges of lamotrigine for patients with epilepsy: A step towards optimizing treatment and increasing cost-effectiveness. Epilepsy research, 176, 106728. 10.1016/j.eplepsyres.2021.106728

19. Evans, B. K., Kustra, R. P., & Hammer, A. E. (2007). Assessment of tolerability in elderly patients: changing to lamotrigine therapy. The American journal of geriatric pharmacotherapy, 5(2), 112–119. 10.1016/j.amjopharm.2007.06.001

20. Brzaković, B. B., Vezmar Kovačević, S. D., Vučićević, K. M., Miljković, B. R., Martinović, Z. J., Pokrajac, M. V., & Prostran, M. Š. (2012). Impact of age, weight and concomitant treatment on lamotrigine pharmacokinetics. Journal of clinical pharmacy and therapeutics, 37(6), 693–697. 10.1111/j.1365-2710.2012.01351.x

21. Williams, M. H., & Broadley, S. A. (2008). SUNCT and SUNA: clinical features and medical treatment. Journal of clinical neuroscience: official journal of the Neurosurgical Society of Australasia, 15(5), 526–534. 10.1016/j.jocn.2006.09.006

22. Lambru, G., Rantell, K., O’Connor, E., Levy, A., Davagnanam, I., Zrinzo, L., & Matharu, M. (2020). Trigeminal neurovascular contact in SUNCT and SUNA: a cross-sectional magnetic resonance study. Brain: a journal of neurology, 143(12), 3619–3628. 10.1093/brain/awaa331

23. Mason, B. N., & Russo, A. F. (2018). Vascular Contributions to Migraine: Time to Revisit?. Frontiers in cellular neuroscience, 12, 233. 10.3389/fncel.2018.00233

24. Shauly, O., Gould, D. J., Sahai-Srivastava, S., & Patel, K. M. (2019). Greater Occipital Nerve Block for the Treatment of Chronic Migraine Headaches: A Systematic Review and Meta-Analysis. Plastic and reconstructive surgery, 144(4), 943–952. 10.1097/PRS.0000000000006059

