## Supplemental file 1 for "Treatment Outcomes and MRI Features of SUNA: A Systematic Review and Meta-Analysis"

**Search Strategy Comprising MeSH Keywords**

*Population of Interest*

((SUNA) OR (Short-Lasting Unilateral Neuralgiform Headache Attacks) OR (Short-Lasting Unilateral Neuralgiform Headache Attacks With Cranial Autonomic Symptoms) OR (Trigeminal Autonomic Cephalalgias))

AND

*Outcomes of Interest*

(Pain, Sudden OR Pain, Sharp OR Pain, Unilateral) OR (Treatment OR Therapy) OR (Magnetic Resonance Imaging OR MRI)
