## Supplemental file 2 for "Treatment Outcomes and MRI Features of SUNA: A Systematic Review and Meta-Analysis"

| PMID | Country | Age | Sex | Brain MRI Findings | Comorbidities | Treatment | Dose | Duration of Follow-up | Clinical Improvement | Complications |
|---|---|---|---|---|---|---|---|---|---|---|
| | | | | | | Lamotrigine | 25 mg/day | 0.3 months | Yes | NA |
| 29484308 | Japan | 57 | F | T2-hyperintense lesions in the left dorsolateral medulla oblongata and left dorsal cervical spinal cord at the C1/2 spine level | Neuromyelitis optica spectrum disorder | Lamotrigine | 75 mg/day | 1 month | Yes | NA |
| | | | | | | Carbamazepine | 400 mg/day | NA | No | NA |
| | | | | | | Indomethacin | 400 mg/day | NA | No | NA |
| | | | | | | Valproate | NA | NA | No | NA |
| | | | | | | | | 0.3 months | Yes | NA |
| 20009409 | Japan | 18 | M | Cortical dysplasia with intrusion of the cortex deep into the precentral sulcus nearly to the lateral ventricle, and the signal level of this lesion was equal to that of the surrounding cortex on T1- and T2-weighted images but it was higher than that in part of the cortex on FLAIR images. | NA | Lomerizine | 10 mg/day | 6 months | Yes | NA |
| | | | | | | Carbamazepine | 1200 mg/day | NA | No | NA |
| | | | | | | Pregabalin | 400 mg/day | NA | No | NA |
| 30482045 | Portugal | 57 | M | Right acoustic neuroma compressing the right facial nerve and a venous developmental anomaly perpendicular to the right facial nerve root entry zone, without lesions affecting the trigeminal nerve. | NA | Lamotrigine | 200 mg/day | NA | Yes | NA |
| | | 29 | M | Vascular loop in close proximity to the right trigeminal nerve | NA | Gabapentin | 900 mg/day | NA | Yes | NA |
| 18513261 | UK | 24 | F | NAD | NA | Carbamazepine | 400 mg/day | 2 months | Yes | NA |
| | | | | | | Carbamazepine | NA | NA | No | NA |
| | | | | | | Ibuprofen | NA | NA | No | NA |
| | | | | | | Gabapentin | 1600 mg/day | NA | No | NA |
| | | | | | | Indomethacin | 75 mg/day | NA | No | NA |
| 21409598 | Spain | 36 | M | Extra-axial lesion at the right cerebellopontine angle, heterogeneous, hyperintense with edges well defined. The lesion produces imprint on the brain stem without signs of infiltration. All these features are suggestive of an epidermoid cyst | Epidermoid cyst | Retrosigmoid craniectomy | NA | NA | Yes | None |
| | | 50 | M | Isodense, thickened dura on T1 and T2, marked dural enhancement in the right cerebral [illegible] | NA | Methylprednisolone | 1000 mg/day | NA | Yes | NA |
| 22077924 | USA | 46 | M | Enhancement of thickened dura in the right hemisphere, MRA and MRV - NAD | NA | Oxcarbazepine | 2400 mg/day | NA | Yes | NA |
| | | | | | | Gabapentin | 2700 mg/day | NA | Yes | Drug resistance |
| | | | | | | Pregabalin | 600 mg/day | NA | Yes | Drug resistance |
| | | | | | | Indomethacin | 150 mg/day | NA | Yes | Drug resistance |
| 17381562 | UK | 32 | F | NA | Stroke | Lamotrigine | 100 mg/day | NA | Yes | None |
| | | | | | | Indomethacin | 75 mg/day | 2 months | No | NA |
| | | | | | | Amitriptyline | NA | NA | No | NA |
| | | | | | | Valproic acid | NA | NA | No | NA |
| | | | | | | Aspirin+caffeine | NA | NA | No | NA |
| | | | | | | Acetaminophen | NA | NA | No | NA |
| | | | | | | Ibuprofen | NA | NA | No | NA |
| | | | | | | Mefenamic acid | NA | NA | No | NA |

| | | | | | | | | | | |
|---|---|---|---|---|---|---|---|---|---|---|
| 15910574 | USA | 11 | F | NAD | NA | Gabapentin | NA | NA | Yes | NA |
| | | | | | | Lamotrigine | 400 mg/day | NA | Yes | NA |
| | | | | | | Gabapentin | 100 mg/day | NA | Yes | NA |
| | | | | | | Baclofen | 40 mg/day | NA | Yes | NA |
| | | | | | | Ice pack application | | NA | Yes | NA |
| | | 62 | M | NAD | Asymptomatic right TMJ disc degeneration; previous facial surgeries: bilateral microvascular decompression, glycerol rhizotomy, uvulopalatopharyngoplasty | Onabotulinumtoxin A | 100 U - left masseter muscle, 100 U - [illegible] | NA | Yes | NA |
| | | | | | | Lamotrigine | 300 mg/day | NA | Yes | NA |
| | | 26 | M | NAD | Trigeminal neuralgia | Gabapentin | 3000 mg/day | NA | Yes | NA |
| | | 57 | F | NAD | Pulmonary neuroendocrine tumor, right TMJ [illegible] | Verapamil | 80 mg/day | NA | Yes | NA |
| 33269943 | USA | 55 | M | T2-hyperintensities with subtle enhancement, several hypointense areas on T1 imaging, [illegible] | Incidental right TMJ disc displacement with [illegible] | Oxcarbazepine | 600 mg/day | NA | Yes | NA |
| | | | | | | Lamotrigine | NA | 1.5 months | Yes (2/6) | NA |
| | | | | | | Gabapentin | NA | 1.5 months | Yes (2/5) | NA |
| | | | | | | Topiramate | NA | 1.5 months | No (4) | NA |
| | | | | | | Carbamazepine | NA | 1.5 months | No (13) | NA |
| | | | | | | Pregabalin | NA | 1.5 months | Yes (1/4) | NA |
| | | | | | | Amitriptyline | NA | 1.5 months | No (4) | NA |
| 38022430 | India | Mean=41.5, n=13 | F=7, M=6 | NA | NA | Duloxetine | NA | 1.5 months | No (4) | NA |
| 33301567 | UK (n=79) | NA | NA | Significantly greater proportion of neurovascular contacts on the symptomatic (74.4%) [illegible] | NA | NA | NA | NA | NA | NA |
| | | | | | | Sumatriptan | NA | NA | Yes (1/9) | NA |
| | | | | | | Oxygen | 12-15 l/min | NA | No (7) | NA |
| | | | | | | Indomethacin | 100 mg | NA | No (6) | NA |
| | | | | | | Lidocaine | NA | NA | Yes (8/9) | NA |
| | | | | | | Dihydroergotamine | NA | NA | No (5) | NA |
| | | | | | | Corticosteroids | NA | NA | No (6) | NA |
| | | | | | | Greater occipital nerve block | NA | NA | Yes (3/4) | NA |
| | | | | | | Lamotrigine | 100-600 mg/day | NA | Yes (5/16) | NA |
| | | | | | | Topiramate | NA | NA | Yes (1/9) | NA |
| | | | | | | Gabapentin | 1800-2400 mg/day | NA | Yes (7/18) | NA |
| | | | | | | Carbamazepine | NA | NA | Yes (4/20) | NA |
| | | | | | | Oxcarbazepine | NA | NA | No (6) | NA |
| | | | | | | Pregabalin | NA | NA | Yes (1/16) | NA |

| | | | | | | | | | | |
|---|---|---|---|---|---|---|---|---|---|---|
| | | | | | | Verapamil | NA | NA | No (5) | NA |
| | | | | | | Valproate | NA | NA | No (4) | NA |
| | | | | | | Beta blocker | NA | NA | No (4) | NA |
| 29096522 | USA, UK | Mean= 45, n=37 | M=18, F=19 | NAD (31/37), vascular loops with trigeminal nerve compression (2/37), NA (4/37) | NA | Tricyclic antidepressants | NA | NA | Yes (3/17) | NA |
| | | | | | | Lamotrigine | NA | NA | No | NA |
| | | | | | | Oxcarbazepine | NA | NA | No | NA |
| | | | | | | Gabapentin | NA | NA | No | NA |
| | | | | | | Greater occipital nerve block | NA | NA | No | NA |
| | | | | | | Amitriptyline | NA | NA | No | NA |
| | | | | | | Topiramate | NA | NA | No | NA |
| | | | | | | Onabotulinum toxin A | NA | NA | No | NA |
| | | 63 | F | NA | NA | Sphenopalatine ganglion Pulsed [illegible] | 100V, 2 times | 30 months | Yes | None |
| | | | | | | Oxcarbazepine | NA | NA | No | NA |
| | | | | | | Onabotulinum toxin A | NA | NA | No | NA |
| | | | | | | Amitriptyline | NA | NA | No | NA |
| | | | | | | Duloxetine | NA | NA | No | NA |
| | | | | | | Valproate | NA | NA | No | NA |
| | | | | | | Lamotrigine | NA | NA | No | NA |
| | | | | | | Carbamazepine | NA | NA | No | NA |
| | | | | | | Gabapentin | NA | NA | No | NA |
| | | | | | | Pregabalin | NA | NA | No | NA |
| | | | | | | Lacosamide | NA | NA | No | NA |
| | | | | | | Greater occipital nerve block | NA | NA | No | NA |
| | | 51 | F | NA | NA | Sphenopalatine ganglion pulsed [illegible] | 100V, single procedure | 18 months | Yes | No |
| | | | | | | Carbamazepine | NA | NA | No | NA |
| | | | | | | Lamotrigine | NA | NA | No | NA |
| | | 47 | F | NA | NA | Sphenopalatine ganglion pulsed [illegible] | 100V, single procedure | 6 months | Yes | None |
| | | | | | | Lamotrigine | NA | NA | No | NA |
| | | | | | | Carbamazepine | NA | NA | No | NA |
| | | | | | | Lacosamide | NA | NA | No | NA |
| | | | | | | Gabapentin | NA | NA | No | NA |

| | | | | | | | | | | |
|---|---|---|---|---|---|---|---|---|---|---|
| | | | | | | Amitriptyline | NA | NA | No | NA |
| | | | | | | Nortriptyline | NA | NA | No | NA |
| | | | | | | Pregabalin | NA | NA | No | NA |
| | | | | | | Topiramate | NA | NA | No | NA |
| 32202666 | UK | 45 | M | NA | NA | Sphenopalatine ganglion pulsed [illegible] | 100V, single procedure | 2 months | No | None |
| | | | | | | Indomethacin | NA | NA | No | NA |
| | | | | | | Lidocaine | NA | NA | Yes | NA |
| | | 50 | F | NAD | Hemiplegic migraine | Greater occipital nerve injection | NA | 0.5 months | Yes | NA |
| | | | | | | Indomethacin | NA | NA | No | NA |
| | | 55 | M | NAD | Hemiplegic migraine | Greater occipital nerve injection | NA | NA | No | NA |
| | | | | | | Indomethacin | NA | NA | No | NA |
| | | | | | | Lamotrigine | 175 mg/day | NA | Yes | NA |
| | | | | | | Topiramate | 200 mg/day | NA | Yes | NA |
| | | | | | | Lidocaine | NA | NA | Yes | NA |
| | | 41 | F | Cerebellar infarct | Hemiplegic migraine | Greater occipital nerve injection | NA | 1.5 months | Yes | NA |
| | | | | | | Indomethacin | NA | NA | No | NA |
| | | | | | | Lamotrigine | 200 mg/day | NA | Yes | NA |
| | | 41 | M | NAD | Hemiplegic migraine | Greater occipital nerve injection | NA | NA | No | NA |
| | | 34 | F | Right frontal cortical dysplasia | Hemiplegic migraine | Greater occipital nerve injection | NA | NA | No | NA |
| | | | | | | Indomethacin | NA | NA | No | NA |
| | | | | | | Lidocaine | NA | NA | Yes | NA |
| | | | | | | Topiramate | 400 mg/day | NA | Yes | NA |
| | | | | | | Oxcarbazepine | 2400 mg/day | NA | Yes | NA |
| 22238357 | UK | 54 | F | NAD | Hemiplegic migraine | Greater occipital nerve injection | NA | 2 months | Yes | NA |
| | | | | | | Sumitriptan | NA | NA | No | NA |
| | | | | | | Oxygen | NA | NA | No | NA |
| 27489179 | Denmark | 16 | M | NAD | NA | Indomethacin, PPI | Initiated with 25 mg/day, increased to [illegible] | 10 months | Yes | NA |
| | | | | | | Carbamazepine | NA | NA | No | NA |
| | | | | | | Lamotrigine | NA | NA | No | NA |
| | | 56 | M | Aberrant loop of superior cerebellar artery | NA | Surgery | NA | 20 months | Yes | Right jaw pain |
| | | | | | | Gabapentin | NA | NA | No | NA |

| | | | | | | | | | | |
|---|---|---|---|---|---|---|---|---|---|---|
| | | | | | | Lamotrigine | NA | NA | No | NA |
| 20462914 | Australia | 48 | F | Aberrant loop of the anterior inferior cerebellar artery and vein | NA | Surgery | NA | 20 months | No | Dural sinus bleeed, hearing [illegible] |
| | | | | | | Nortryptyline | 50 mg/day | NA | No | NA |
| | | | | | | Gabapentin | 1800 mg/day | NA | No | NA |
| | | | | | | Topiramate | 75 mg/day | NA | No | NA |
| 23565730 | UK | 64 | F | NAD | Red ear syndrome | Lamotrigine | 400 mg/day | NA | Yes | NA |
| | | | | | | Lamotrigine | 300 mg/day | NA | No | NA |
| | | | | | | Topiramate | 700 mg/day | NA | No | NA |
| | | | | | | Gabapentin | 2400 mg/day | NA | No | NA |
| | | | | | | Pregabalin | 350 mg/day | NA | No | NA |
| | | | | | | Carbamazepine | 1600 mg/day | NA | No | NA |
| | | | | | | Mexiletine | 1200 mg/day | NA | No | NA |
| | | | | | | Melatonin | 9 mg/day | NA | No | NA |
| | | | | | | Amitriptyline | 25 mg/day | NA | No | NA |
| | | | | | | Tizanidine | NA | NA | No | NA |
| | | | | | | Indomethacin | 225 mg/day | 0.75 month | No | NA |
| | | | | | | Oxygen | NA | NA | No | NA |
| | | | | | | Sumatriptan | 6 mg/day | NA | No | NA |
| | | | | | | Lidocaine | NA | 0 months | Yes | NA |
| | | | | | | Greater occipital nerve block | NA | | Yes | NA |
| | | 61 | F | NAD | NA | Occipital nerve stimulation | NA | 55 months | Yes | Battery discharged after 34 [illegible] |
| | | | | | | Lamotrigine | 250 mg/day | NA | No | NA |
| | | | | | | Topiramate | 150 mg/day | NA | No | NA |
| | | | | | | Gabapentin | 3600 mg/day | NA | No | NA |
| | | | | | | Pregabalin | 600 mg/day | NA | No | NA |
| | | | | | | Oxcarbazepine | 1500 mg/day | NA | No | NA |
| | | | | | | Melatonin | 15 mg/day | NA | No | NA |
| | | | | | | Amitriptyline | 50 mg/day | NA | No | NA |
| | | | | | | Sodium valproate | 1000 mg/day | NA | No | NA |
| | | | | | | Indomethacin | 100 mg/day | NA | No | NA |
| | | | | | | Oxygen | NA | NA | No | NA |

| | | | | | | | | | | |
|---|---|---|---|---|---|---|---|---|---|---|
| | | | | | | Sumatriptan | 6 mg/day | NA | No | NA |
| | | | | | | Lidocaine | NA | NA | Yes | NA |
| | | 34 | F | NAD | NA | Occipital nerve stimulation | NA | 28 months | Yes | Battery discharged after 25 [illegible] |
| | | | | | | Lamotrigine | 200 mg/day | NA | No | NA |
| | | | | | | Topiramate | 200 mg/day | NA | No | NA |
| | | | | | | Gabapentin | 3600 mg/day | NA | No | NA |
| | | | | | | Pregabalin | 400 mg/day | NA | No | NA |
| | | | | | | Oxcarbazepine | 1200 mg/day | NA | No | NA |
| | | | | | | Melatonin | 12 mg/day | NA | No | NA |
| | | | | | | Amitriptyline | 50 mg/day | NA | No | NA |
| | | | | | | Pizotifen | 3 mg/day | NA | No | NA |
| | | | | | | Indomethacin | 100 mg/day | NA | No | NA |
| | | | | | | Oxygen | NA | NA | No | NA |
| | | | | | | Sumatriptan | 6 mg/day | NA | No | NA |
| | | | | | | Lidocaine infusion | NA | 0.3 months | Yes | NA |
| | | | | | | Greater occipital nerve block | NA | NA | No | NA |
| 24452643 | UK | 33 | F | NAD | NA | Occipital nerve stimulation | NA | 28 months | Yes | Moderate neck stiffness. [illegible] |
| | | | | | | Oxygen | NA | NA | Yes (1/2) | NA |
| | | | | | | Triptan | NA | NA | No (1) | NA |
| | | | | | | Indomethacin | NA | NA | No (2) | NA |
| | | | | | | Carbamezapine | NA | NA | Yes (1/3) | NA |
| | | | | | | Corticosteroid | NA | NA | Yes (1/3) | NA |
| | | | | | | Lamotrigine | NA | NA | No (2) | NA |
| | | | | | | Topiramate | NA | NA | Yes (1) | NA |
| | | | | | | Gabapentin | NA | NA | Yes (1/3) | NA |
| | | | | | | Pregabalin | NA | NA | Yes (2/3) | NA |
| | | | | | | Verapamil | NA | NA | No (1) | NA |
| | | | | | | Traditional Chinese medicine | NA | NA | Yes (1/5) | NA |
| | | | | | | Anesthetic blockade | NA | NA | No (1) | NA |
| 36289482 | China | Mean=48.6, n=31 | F=16, M=15 | NA | NA | Acupuncture | NA | NA | No (2) | NA |
| | | | | | | Lamotrigine | Mean=348.7 mg (150-600 mg/day) | NA | Yes (43/60) | NA |

| | | | | | | | | | | |
|---|---|---|---|---|---|---|---|---|---|---|
| | | | | | | Carbamazepine | Mean=750 mg (200-2000 mg/day) | NA | Yes (26/43) | Side effects |
| | | | | | | Gabapentin | Mean=1940.7 mg (200-4800 mg/day) | NA | Yes (10/30) | Side effects |
| | | | | | | Topiramate | Mean=285 mg (100-400 mg/day) | NA | Yes (10/31) | Side effects |
| | | | | | | Pregabalin | Mean=355.4 mg (75-600 mg/day) | NA | Yes (8/29) | Side effects |
| | | | | | | Oxcarbazepine | Mean=1591.7 mg (600-3600 mg/day) | NA | Yes (25/34) | Side effects |
| | | | | | | Duloxetine | Mean=90mg (30-120mg/day) | NA | Yes (6/17) | Tolerability issues |
| | | | | | | Mexiletine | Mean=916.7 mg (400-1200 mg/day) | NA | Yes (3/7) | Side effects |
| | | | | | | Lacosamide | Mean=120 mg (200-400 mg/day) | NA | Yes (1/5) | Not tolerated by 2 patients |
| | | | | | | Lidocaine | NA | 3 months | Yes (21/23) | Cognitive slowing, paranoid [illegible] |
| 33361408 | UK | Mean= 43.7, n=76 | M= 23, F= 53 | NA | NA | Greater occipital nerve block | NA | 1.5 months | Yes (14/31) | Worsening headache in non [illegible] |
| **Abbreviations Used** | | | | | | | | | | |
| NA | data not available | | | | | | | | | |
| UA | not applicable | | | | | | | | | |
| NAD | no abnormality detected | | | | | | | | | |
