## Supplemental file 3 for "Treatment Outcomes and MRI Features of SUNA: A Systematic Review and Meta-Analysis"

| Study | Treatment Modality | Frequency (per day) | | Duration (minutes) | | |
|---|---|---|---|---|---|---|
| | | Before Drug | After Drug | Before Drug | After Drug | |
| | indomethacin | 30 | 30 | NA | NA | |
| | amitryptline | 30 | 30 | NA | NA | |
| | valproate | 30 | 30 | NA | NA | |
| | aspirin + caffeine | 30 | 30 | NA | NA | |
| | acetaminophen | 30 | 30 | NA | NA | |
| | ibuprofen | 30 | 30 | NA | NA | |
| Volcy, et al., 2004 | mefenamic acid | 30 | 30 | NA | NA | |
| | gabapentin | 10 | 0.14 | 1 | NA | patient 1 |
| Jacob, et al., 2008 | carabmazepine | 8 | 2 | 3 | NA | patient 2 |
| | valproate | 5 | 5 | 5 | 5 | |
| | carabmazepine | 5 | 5 | 5 | 5 | |
| | loxoprofen | 5 | 5 | 5 | 5 | |
| | diclofenac | 5 | 5 | 5 | 5 | |
| | indomethacin | 5 | 5 | 5 | 5 | |
| Tada, et al., 2009 | lomerizine | 5 | 0.28 | 5 | 2 | |
| Caballero, et al., 2011 | retrosigmoid craniectomy | 12 | 0 | 0.5 | 0 | |
| | methylprednisolone | 15 | 0 | 3 | 0 | patient 1 |
| Chan, 2011 | oxcarbazepine | 12 | NA | 5 | NA | patient 2 |
| | nortriptyline | 200 | 200 | 2 | 2 | |
| | gabapentin | 200 | 200 | 2 | 2 | |
| | topiramate | 200 | 200 | 2 | 2 | |
| Lambru, et al., 2013 | lamotrigine | 200 | 80 | 2 | NA | |
| | lamotrigine | 52 | 52 | 10 | 10 | |
| | topiramate | 52 | 52 | 10 | 10 | |
| | gabapentin | 52 | 52 | 10 | 10 | |
| | pregabalin | 52 | 52 | 10 | 10 | |
| | carbamazepine | 52 | 52 | 10 | 10 | |
| | oxcarbazepine | 52 | 52 | 10 | 10 | |
| | mexilitine | 52 | 52 | 10 | 10 | |
| | melatonin | 52 | 52 | 10 | 10 | |

| | | | | | | |
|---|---|---|---|---|---|---|
| | amitryptline | 52 | 52 | 10 | 10 | |
| | valproate | 52 | 52 | 10 | 10 | |
| | occipital nerve stimulation | 52 | 0 | 10 | 0 | patient 1 |
| | lamotrigine | 42 | 42 | 30 | 30 | |
| | topiramate | 42 | 42 | 30 | 30 | |
| | gabapentin | 42 | 42 | 30 | 30 | |
| | pregabalin | 42 | 42 | 30 | 30 | |
| | carbamazepine | 42 | 42 | 30 | 30 | |
| | oxcarbazepine | 42 | 42 | 30 | 30 | |
| | mexilitine | 42 | 42 | 30 | 30 | |
| | melatonin | 42 | 42 | 30 | 30 | |
| | amitryptline | 42 | 42 | 30 | 30 | |
| | valproate | 42 | 42 | 30 | 30 | |
| | occipital nerve stimulation | 42 | 12 | 30 | 0.3 | patient 2 |
| | lamotrigine | 154 | 154 | 10 | 10 | |
| | topiramate | 154 | 154 | 10 | 10 | |
| | gabapentin | 154 | 154 | 10 | 10 | |
| | pregabalin | 154 | 154 | 10 | 10 | |
| | carbamazepine | 154 | 154 | 10 | 10 | |
| | oxcarbazepine | 154 | 154 | 10 | 10 | |
| | mexilitine | 154 | 154 | 10 | 10 | |
| | melatonin | 154 | 154 | 10 | 10 | |
| | amitryptline | 154 | 154 | 10 | 10 | |
| | valproate | 154 | 154 | 10 | 10 | |
| Lambru, et al., 2014 | occipital nerve stimulation | 154 | 1.7 | 10 | 4 | patient 3 |
| | sumatriptan | 60 | 60 | 0.1 | NA | |
| | oxygen | 60 | 60 | 0.1 | NA | |
| Cvetkovic, et al., 2017 | indomethacin | 60 | 8 | 0.1 | NA | |
| Mizuno, et al., 2018 | lamotrigine | 20 | 0 | 3 | 0 | |
| | carbamazepine | 50 | 50 | 1 | 1 | |
| | pregabalin | 50 | 50 | 1 | 1 | |

| | | | | | |
|---|---|---|---|---|---|
| Sardoeira, et al., 2019 | lamotrigine | 50 | 0 | 1 | 0 |
| NA | data not available | | | | |
