## Supplemental file 4 for "Treatment Outcomes and MRI Features of SUNA: A Systematic Review and Meta-Analysis"

**Funnel Plots Showing the Publication Bias for Studies Included in the Meta-Analysis on the Proportion of Patients in Which a Treatment Modality was Clinically Effective**

1. Lamotrigine

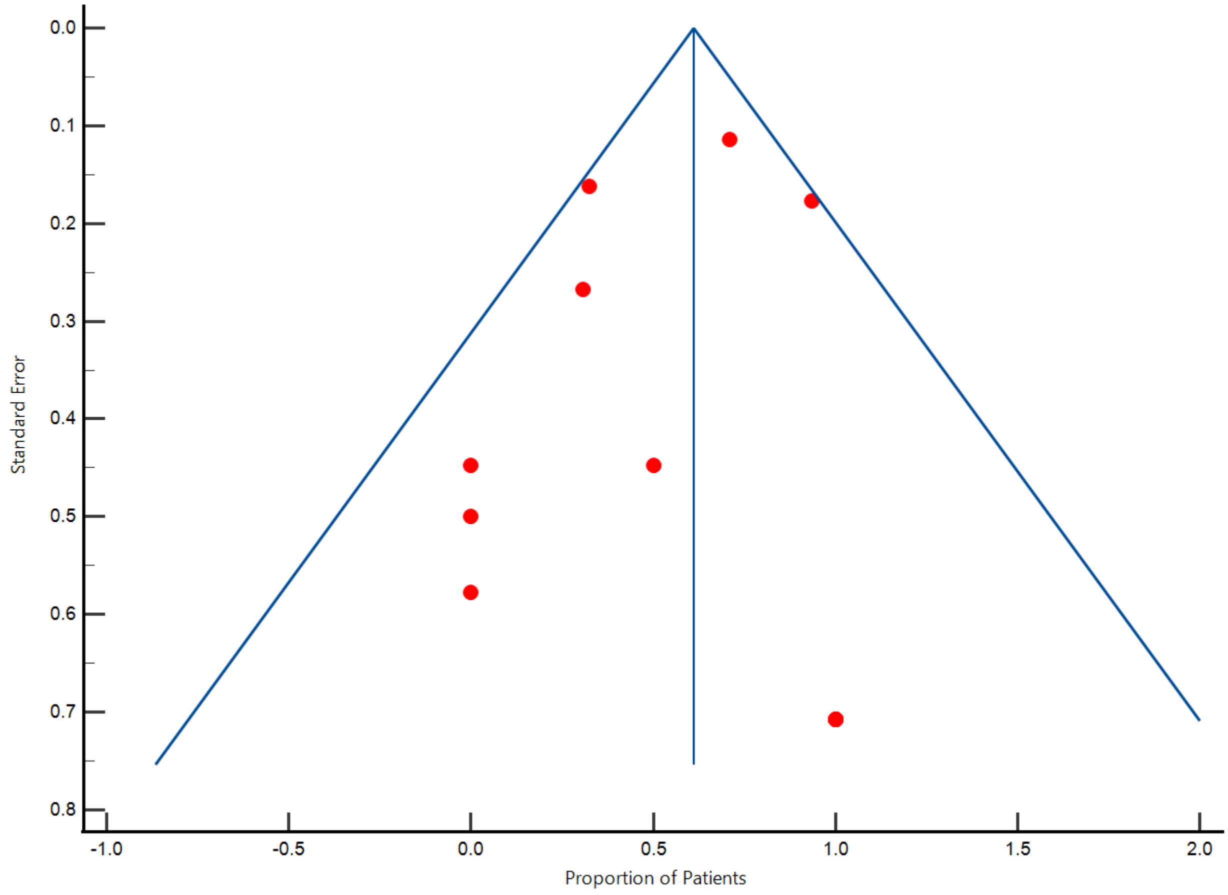

2. Carbamazepine

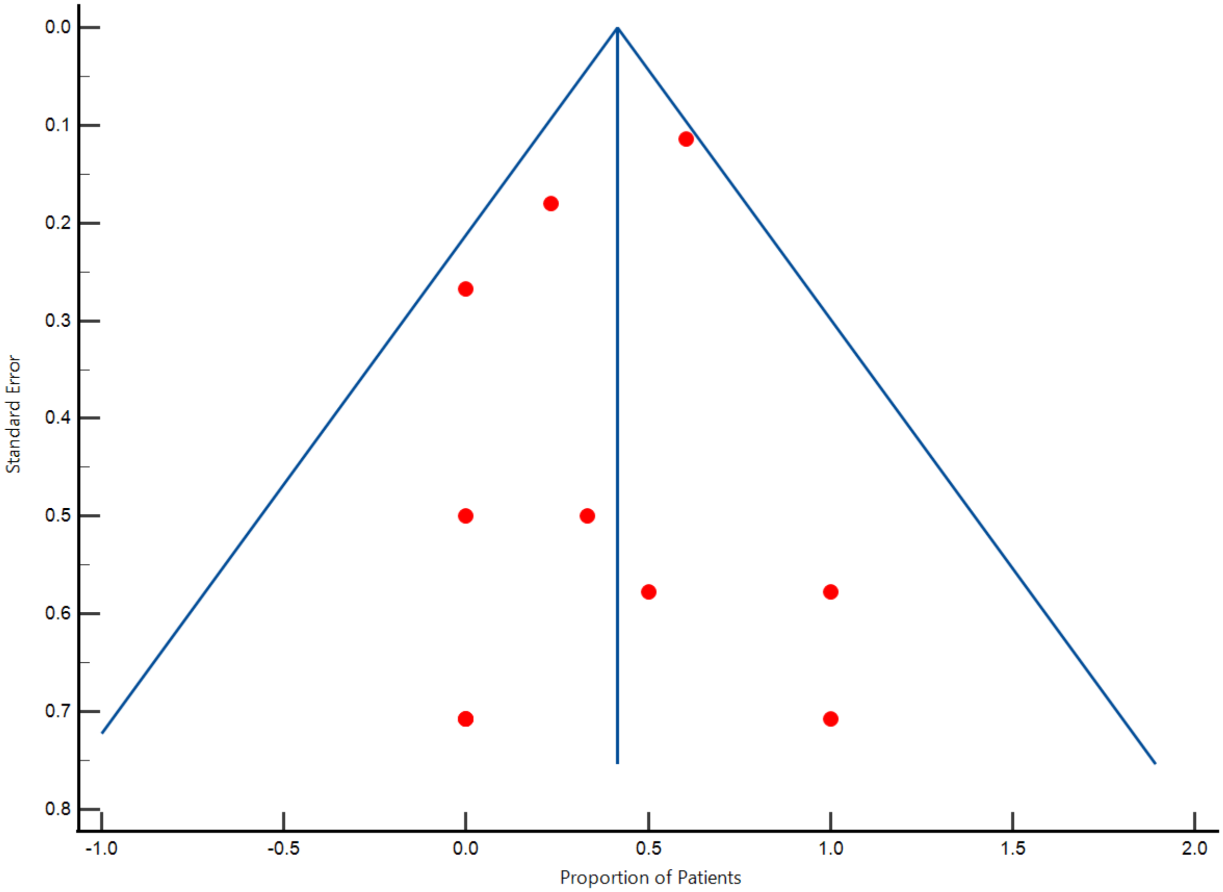

3. Gabapentin

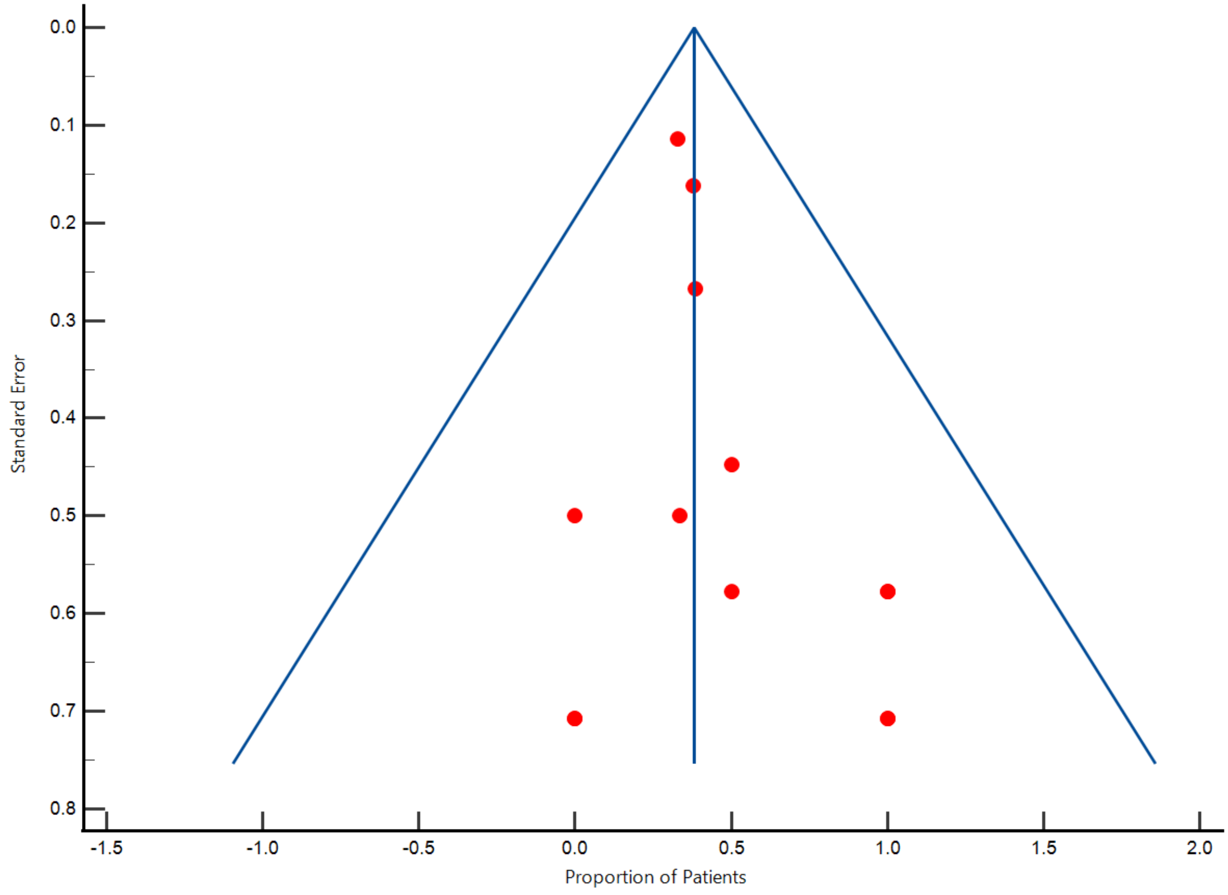

4. Pregabalin

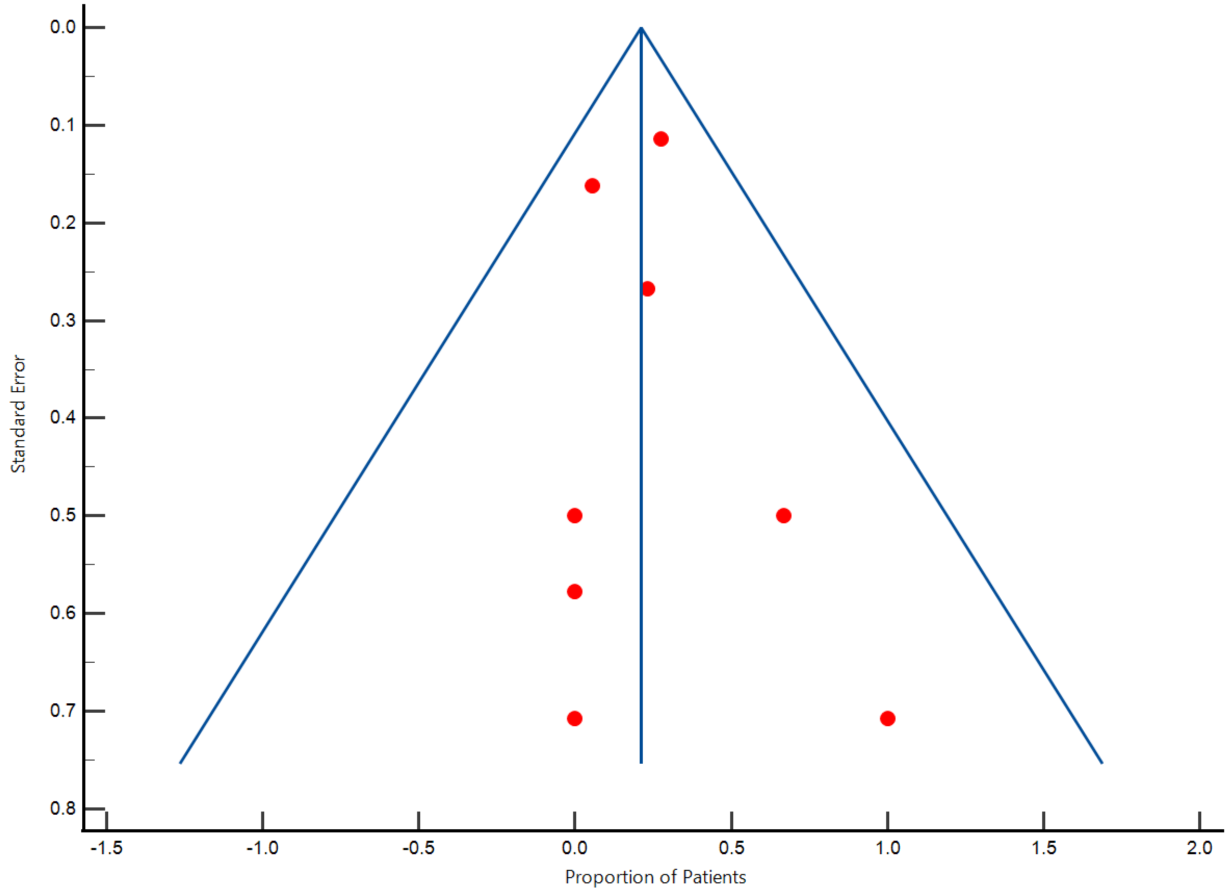

5. Oxcarbazepine

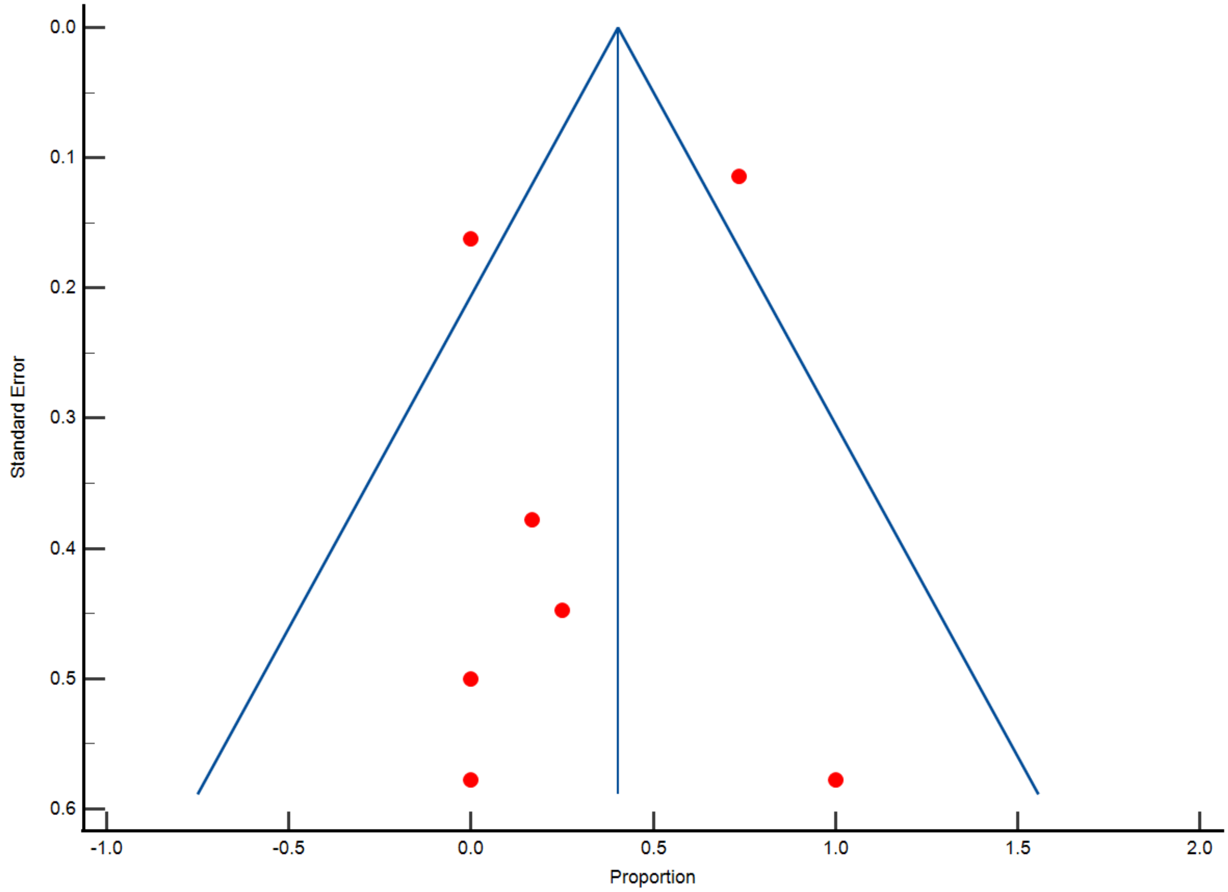

6. Greater Occipital Nerve (GON) Block

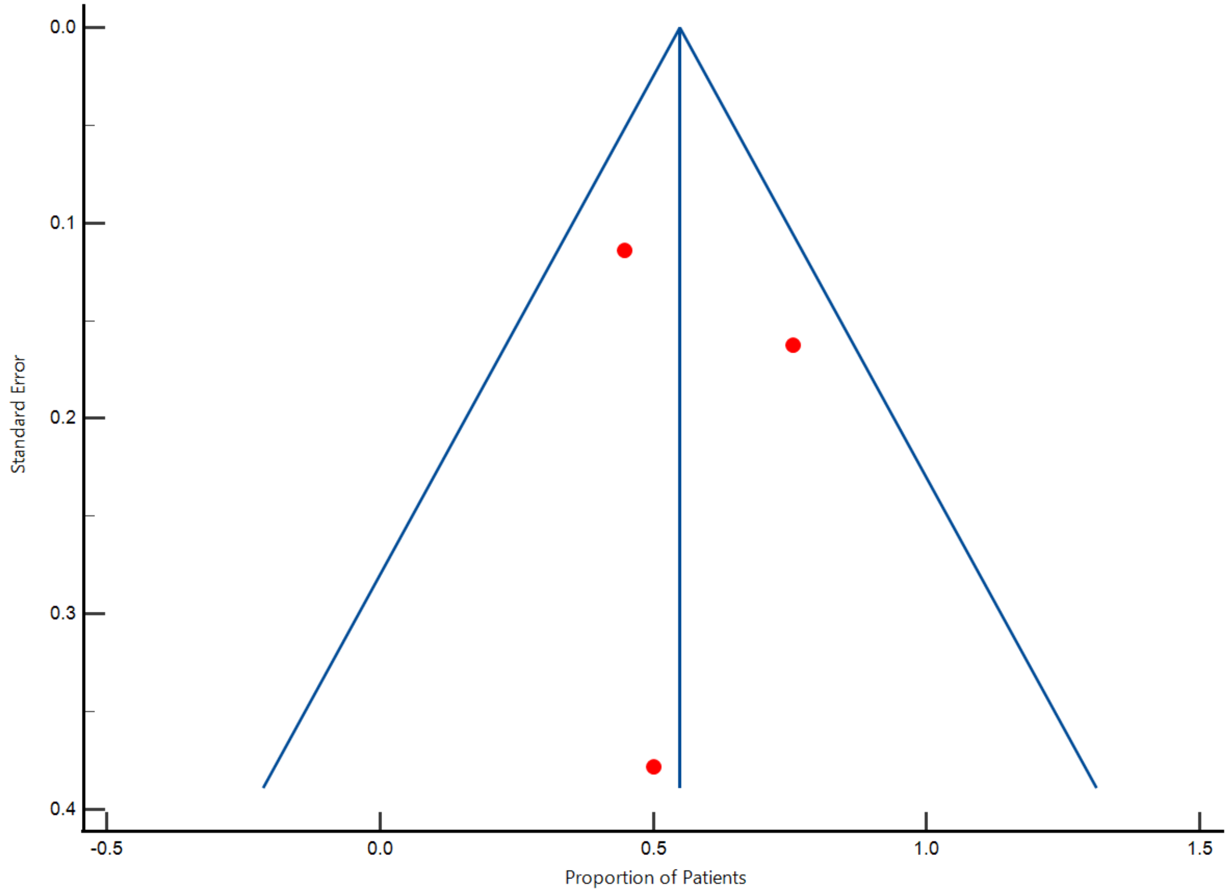

7. Lidocaine

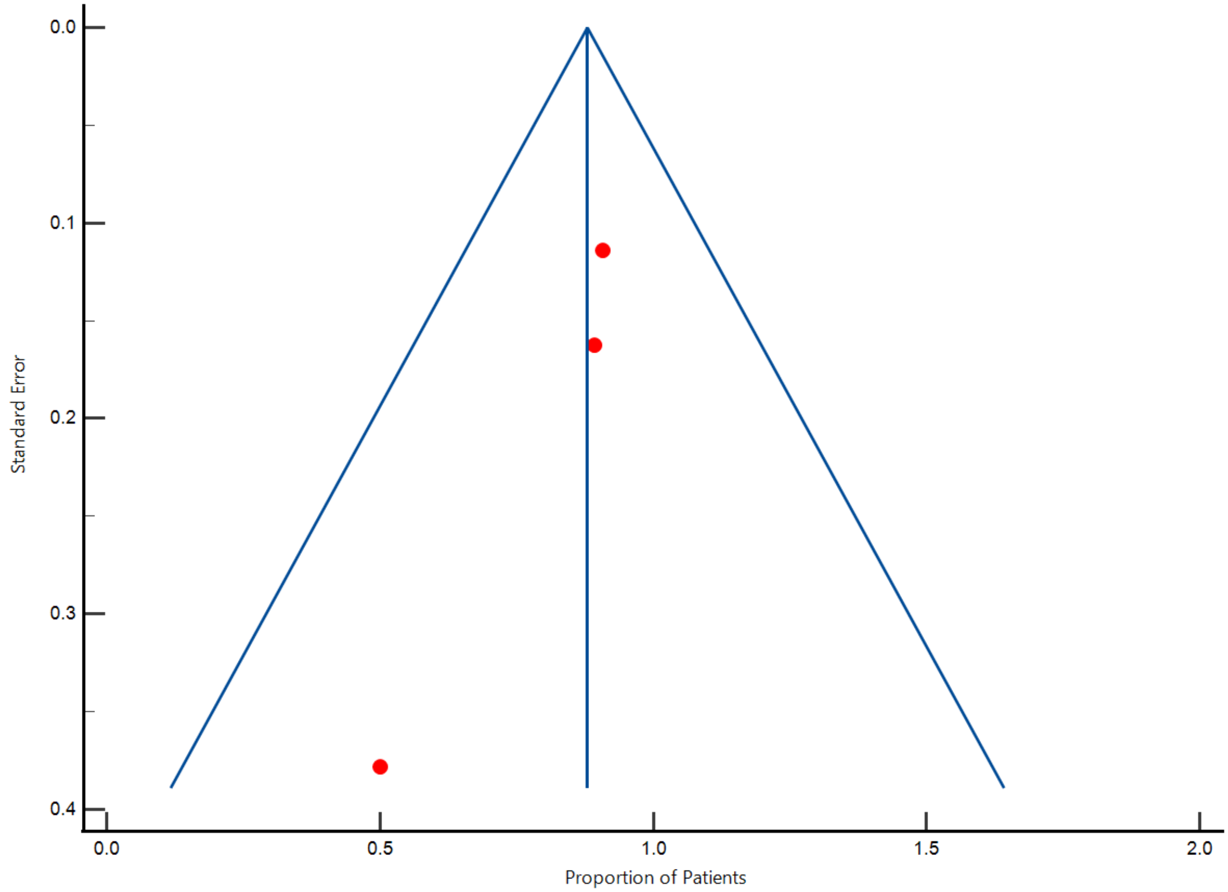

0.0
0.1
0.2
0.3
0.4
Standard Error
0.0
0.5
1.0
1.5
2.0
Proportion of Patients
