## Supplemental file 6 for "Treatment Outcomes and MRI Features of SUNA: A Systematic Review and Meta-Analysis"

| Study | Total score | Selection | | | | Comparability | Outcome | |
|---|---|---|---|---|---|---|---|---|
| | | Representativeness of the sample | Sample size | Non-respondents/loss to follow up | Exposure ascertainment | | Assessment of outcome | Statistical test |
| Mizuno, et al., 2018 | 6 | * | | * | * | * | ** | |
| Tada, et al., 2009 | 6 | * | | * | * | * | ** | |
| Sardoeira, et al., 2019 | 7 | * | | * | * | ** | ** | |
| Jacob, et al., 2008 | 7 | * | | * | * | ** | ** | |
| Caballero, et al., 2011 | 7 | * | | * | * | ** | ** | |
| Chan, 2011 | 7 | * | | * | * | ** | ** | |
| Jacob, et al., 2006 | 6 | * | | * | * | * | ** | |
| Volcy, et al., 2004 | 7 | * | | * | * | ** | ** | |
| R Groenke, et al., 2021 | 8 | * | * | * | * | ** | ** | |
| Prakash, et al., 2023 | 8 | * | * | * | * | ** | ** | |
| Lambru, et al., 2020 | 9 | * | * | * | * | ** | ** | * |
| Weng, et al., 2018 | 9 | * | * | * | * | ** | ** | * |
| Ornello, et al., 2020 | 8 | * | * | * | * | ** | ** | |
| Lambru, et al., 2012 | 8 | * | * | * | * | ** | ** | |
| Cvetkvić, et al., 2017 | 7 | * | | * | * | ** | ** | |
| Williams, et al., 2015 | 9 | * | * | * | * | ** | ** | * |
| Lambru, et al., 2013 | 7 | * | | * | * | ** | ** | |
| Lambru, et al., 2014 | 8 | * | * | * | * | ** | ** | |
| Zhang, et al., 2022 | 9 | * | * | * | * | ** | ** | * |
| Lambru, et al., 2020 | 9 | * | * | * | * | ** | ** | * |
